# Why promising drugs are shelved and barriers and facilitators to re-purposing them: A systematic literature review

**DOI:** 10.1101/2021.09.28.21264254

**Authors:** Nithya Krishnamurthy, Alyssa A. Grimshaw, Sydney A. Axson, Sung Hee Choe, Jennifer E. Miller

## Abstract

**Background:** Despite enthusiasm on the role of repurposing in drug development, enhanced by the Covid-19 pandemic with the FDA granting emergency use authorization of several repurposed drugs to treat Covid-19, there remain knowledge gaps on why pharmaceutical companies abandon the development of promising drug candidates as well as facilitators and barriers to moving them back into development, a process often referred to as drug repurposing.

**Method:** This systematic literature review used a combination of controlled vocabulary and free text terms related to the de-prioritization, shelving, abandonment and repurposing of promising experimental drugs unapproved by the FDA for any indication, to search ABI/Informa, Academic Search Premier, Business Source Complete, Cochrane Library, EconLit, Google Scholar, Ovid Embase, Ovid Medline, Pubmed, Scopus, and Web of Science Core Collection databases. The main outcomes of interest were the characteristics and reasons for the phenomenon of companies deprioritizing or abandoning development of promising drugs, facilitators and successful examples of advancing development of promising abandoned or deprioritized drugs (often referred to as drug repositioning or re-purposing), and barriers to advancing development of promising abandoned or de-prioritized drugs. Study inclusion was not limited by publication date or type. Data extracted included article type, article title, journal title, first author, publication date, extraction and analysis of terminology used to describe abandoned investigational drugs and moving them back into research and development, reason(s) and methods for drug de-prioritization or abandonment, conditions treated, examples of deprioritized or repurposed drugs, as well as barriers and facilitators to drug repurposing. Risk of bias was not performed due to the varying study designs included in this study. Instead, Oxford Centre for Evidence-Based Medicine: Levels of Evidence was used to grade the level of evidence included in this study.

**Results:** We identified 11,814 articles, screening 5,976 for relevance, finding 437 eligible for full text review, 115 of which were included in full analysis. Most articles (66%, 76/115) provided reasons for why drug development may be abandoned, with lack of efficacy, or superiority to other therapies, for the studied indication (n=59), strategic business reasons (n=35), safety problems (n=28), research design decisions (n=12), the complex nature of a studied disease or drug (n=7) and regulatory bodies requiring more information (n=2) among the top. Inadequate resources (n=42) including expertise (n=11), intellectual property challenges (n=26), poor data access (n=20), and uncertainty about the value of repurposing (n=13) along with liability risks (n=5) are key barriers to repurposing. The most common facilitators of drug repurposing were multi-partner collaborations (n=38), access to comprehensive compound databases and corresponding screening tools (n=32), regulatory modifications (n=5) and tax incentives (n=2).

**Conclusion:** More research is needed on the current value of repurposing in drug development, as there remain uncertainties, as well as on how to better facilitate access to resources to support it, where valuable. Financial barriers, insufficient staffing focused on out-licensing shelved products, and challenges negotiating IP agreements in multi-partner collaborations were discussed as barriers to repurposing without clear solutions, suggesting more research is needed in this area.

**Registration:** The protocol was registered on Open Science Framework (https://osf.io/f634k/) as it was not eligible for registration on PROSPERO.

## INTRODUCTION

Drug repurposing, defined as researching new indications for already approved drugs or advancing previously studied but unapproved drugs, is as a core approach in drug development. Some reports state that about 30-40% of new drugs and biologics approved by the US FDA between 2007 and 2009 can be considered repurposed or repositioned products [1]. Similarly, a study found that 35% of transformative drugs approved by the FDA between 1984 and 2009, defined as drugs that were both innovative and had groundbreaking effects on patient care, were repurposed products [2].

Many experts claim re-purposing drugs can be faster, cheaper, less risky and carry higher success rates than traditional drug development approaches, primarily because researchers can bypass earlier stages of development that establish drug safety, as they have already been completed [3]. However, exactly how much time, risk and money are saved can be unclear, with some conflicting evidence.

Some reviews, for example, state about 30% of repurposing efforts are successful, that is result in a product approved for marketing, in comparison to about 10% for new drug applications more generally [4]. However, others conclude contradictorily that repurposed agents do not necessarily succeed more often than new agents, with efficacy typically being the limiting factor rather than safety [5]. In terms of time and cost savings, reports indicate de novo drug discovery and development can be a 10 to 17 year process, in contrast to repurposed drugs which are generally approved sooner, within 3 five 12□years, and at about half the cost [6] [7].

Repurposing is receiving renewed attention during the Covid-19 pandemic [7]. Within six months of the start of the pandemic, the US Food and Drug Administration (FDA) granted emergency use authorization for several repurposed drugs to treat Covid-19, such as remdesivir, originally developed as an antiviral but not previously approved for any indications. Since the start of the pandemic, hundreds of clinical trials of repurposed molecules have been initiated for COVID-19, with lackluster results.

Despite the enthusiasm around drug repurposing, there has been no systematic literature review on why pharmaceutical companies de-prioritize or abandon promising drug candidates in the first place, coupled with an identification of the facilitators and barriers for repurposing promising compounds. Accordingly, this study aims to systematically review the literature to identify the root causes associated with companies shelving development of seemingly promising drug candidates unapproved by the FDA for any indication, as well as obstacles and facilitators for moving them off the shelf and back into development, a process often referred to as drug repurposing.

## METHODS

The Preferred Reporting Items for Systematic Reviews and Meta-analyses (PRISMA) statement for reporting was used for this study (Supplementary Table 1). The protocol was registered on Open Science Framework (https://osf.io/f634k/) as it was not eligible for registration on PROSPERO.

### Search strategy

A systematic search of the literature was conducted by a medical librarian (AAG) in ABI/Informa, Academic Search Premier, Business Source Complete, Cochrane Library, EconLit, Google Scholar, Ovid Embase, Ovid Medline, Pubmed, Scopus, and Web of Science Core Collection databases to find relevant articles published from inception of each database to April 16, 2020. Databases were searched using a combination of controlled vocabulary and free text terms related to the de-prioritization, shelving, abandonment, and repurposing of promising experimental drugs unapproved by the FDA for any indication. The search was peer-reviewed by a second medical librarian using PRESS (Peer Review of Electronic Search Strategies). Details of the full search strategy are listed in Supplementary Table 2.

### Study Selection

Citations from all databases were imported in an Endnote x9 library (Clarivate Analytics, Philadelphia, PA), where duplicates were removed. The de-duplicated results were imported into Covidence v2627 (Covidence, Melbourne, Victoria, Australia) for screening and data extraction. Two independent trained screeners performed a title and abstract review; disagreements were resolved through discussion (SA, NK, JM). The full text of the resulting papers was then reviewed for inclusion by two independent screeners with disagreements also resolved through discussion. The main outcomes of interest were the characteristics of and reasons for the phenomenon of companies deprioritizing or abandoning development of promising drugs, facilitators and successful examples of advancing development of promising abandoned or deprioritized drugs (often referred to as drug repositioning or re-purposing), and barriers to advancing development of promising abandoned or de-prioritized drugs. Study inclusion was not limited by publication date or type. Commentaries, editorials, expert opinions, and perspective pieces were included. Book chapters, conference abstracts, animal studies, dissertations, and papers not available in English were excluded.

### Data Extraction and Analysis

Two reviewers (SA, NK) independently extracted data using Qualtrics software (Qualtrics, Provo, UT). JM performed data extraction on 20% of the final sample, selected at random to verify data reliability. Descriptive analysis was performed by NK, SA and JM. Data extracted included article type, article title, journal title, first author, publication date, extraction and analysis of terminology used to describe abandoned investigational drugs and moving them back into research and development, reason(s) and methods for drug de-prioritization or abandonment, conditions treated, examples of deprioritized or repurposed drugs, as well as barriers and facilitators to drug repurposing. Risk of bias was not performed due to the varying study designs included in this study. Instead, Oxford Centre for Evidence-Based Medicine: Levels of Evidence was used to grade the level of evidence included in this study.

## RESULTS

### Sample characteristics

We identified 11,814 articles through our literature review, 5,838 of which were duplicates. After de-duplicating the sample, we screened 5,976 articles for relevance, finding 437 eligible for full text review, 115 of which were included in our full analysis (See Figure 1 and Supplement Table 3). Of these 115 publications, 18 were expert opinions/editorials, 25 were reviews, 32 were articles, 31 news articles, and 9 were other formats such as commentaries, technical reports, viewpoints, and correspondence (Supplement Table 4).

**Figure 1.**
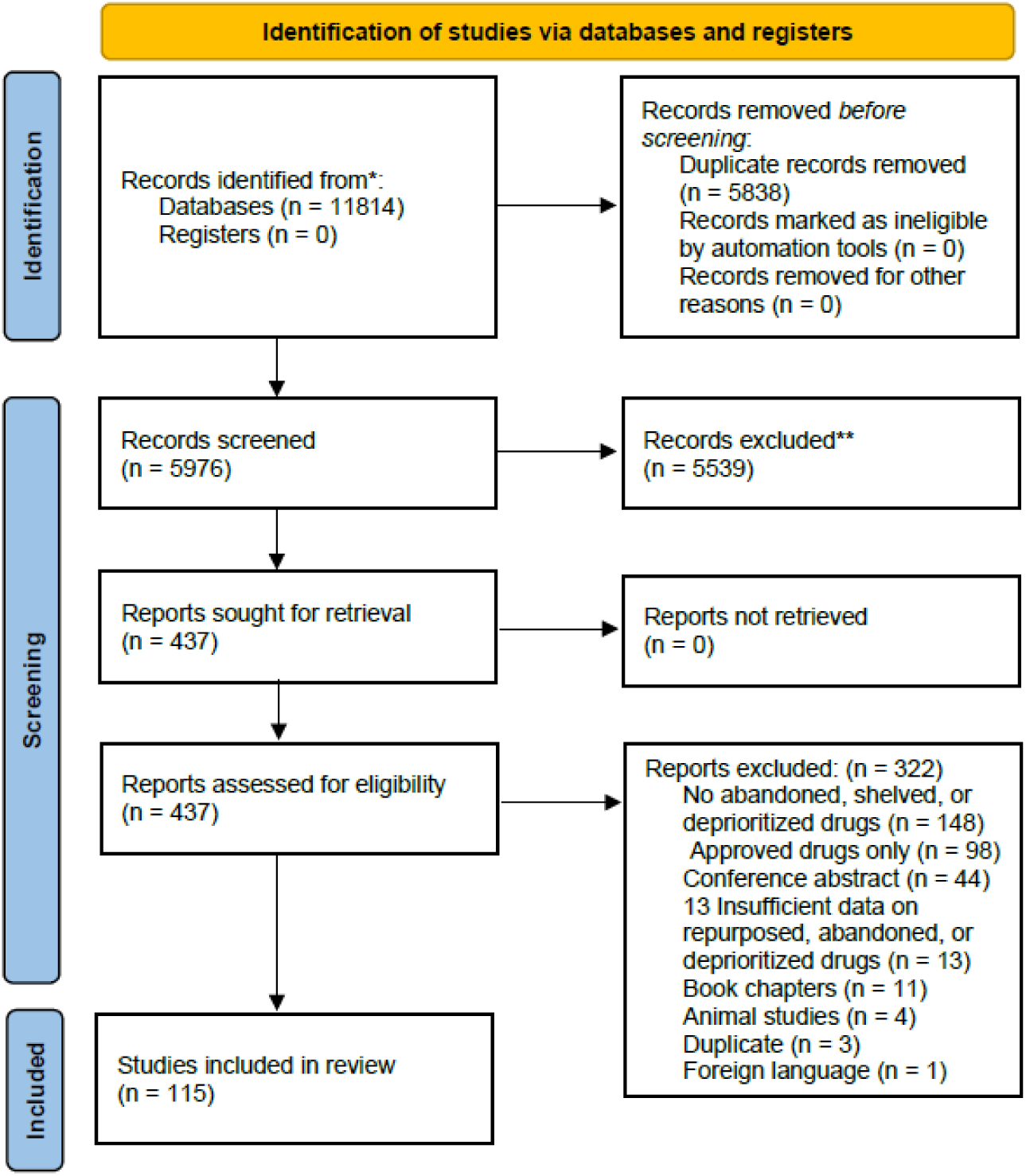
Prisma 2009 Flow Diagram

Sixty-six percent of these articles (76) presented reasons why promising drugs are abandoned (i.e., because a drug is projected not to be a blockbuster or to be less commercially viable than another portfolio drug) and 43% (49) discussed barriers and 63% (72) facilitators for repurposing. The number of articles published on drug repurposing and abandonment has grown over time, from 6 published before 2004, 12 in the period 2005 to 2009, 41 in 2010 to 2014, 51 in 2015-2019, and 5 in 2020, through April 16, 2020 (See Figure 2).

**Figure 2.**
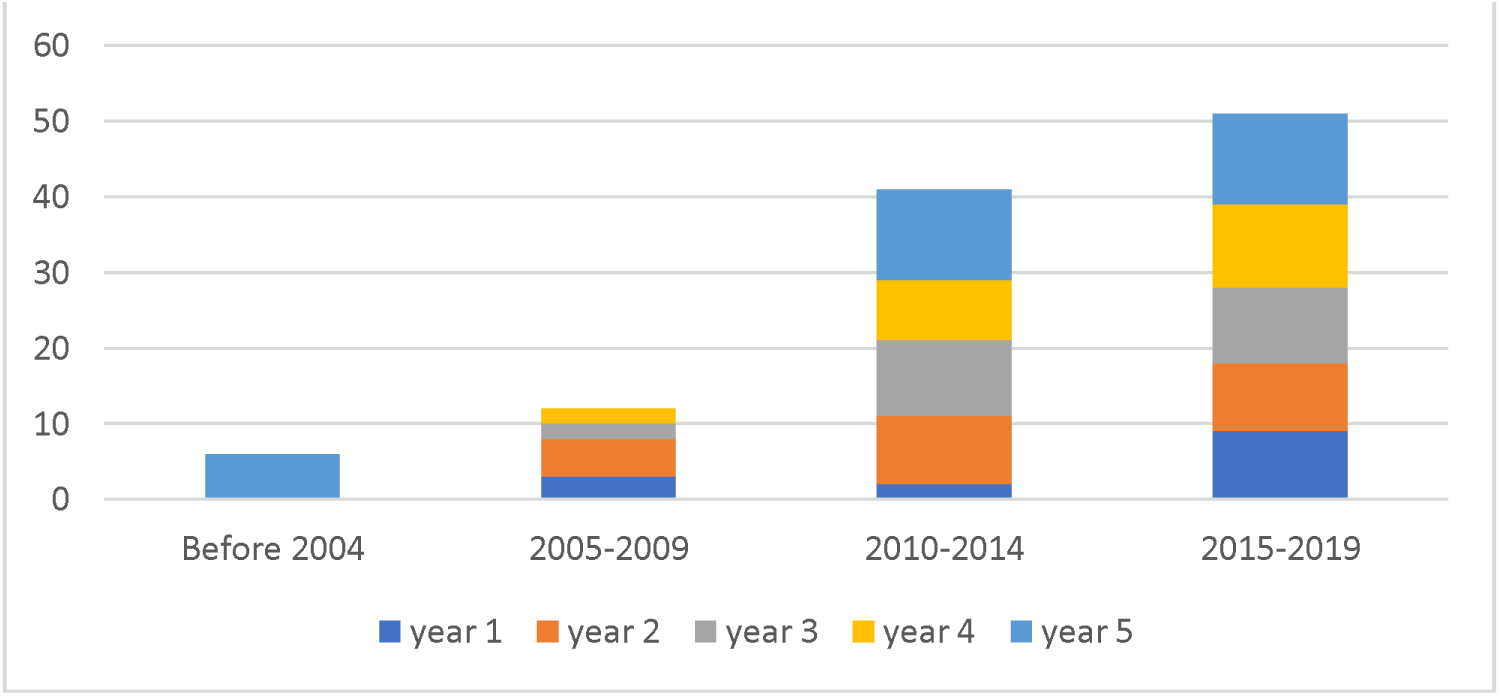
No. papers published per year on the abandonment and repurposing of promising drugs through 2020* **\*** Five papers were also published from Jan 1, 2020 to April 16, 2020.

### Concept definition

Drug candidates pursued by developers not reaching a commercial market are commonly referred to as failed (n=26), abandoned (n=23), discontinued (n=7), shelved (n=8) or deprioritized (n=5), hereafter referred to simply as abandoned. Re-starting investigation of an abandoned drug is commonly referred to as drug repurposing (n=47), repositioning (n=41), reprofiling (n=12), rescue (n=12) and re-tasking (n=5) in the literature. Several articles (n=7) describe how these terms are often used interchangeably in the literature and policy efforts, without consistent definitions, a finding confirmed by our analysis. However, some articles (n=14) distinguish drug repurposing from repositioning, generally referring to repurposing as researching new indications for approved drugs already on the market (i.e., investigating applications for entirely new therapeutic areas), in contrast to repositioning which develops previously studied but unapproved active pharmacological ingredients. Of the articles using repurposing as the primary term and providing an operational definition, 13 stated repurposing applied to both approved and unapproved compounds.

### Reasons drugs are abandoned

Most articles (76/115, 66%) included a discussion on the reasons why a drug candidate’s development may be abandoned, with lack of efficacy for the studied indication (n=59), strategic business reasons by the sponsor (n=35), drug safety problems (n=28), and research design decisions (n=12) being the most commonly discussed reasons. Other cited reasons included the complex nature of the studied disease or drug (n=7) and regulatory bodies requiring more information (n=2) (See Figure 3). Below we go into more detail about some of these reasons.

**Figure 3.**
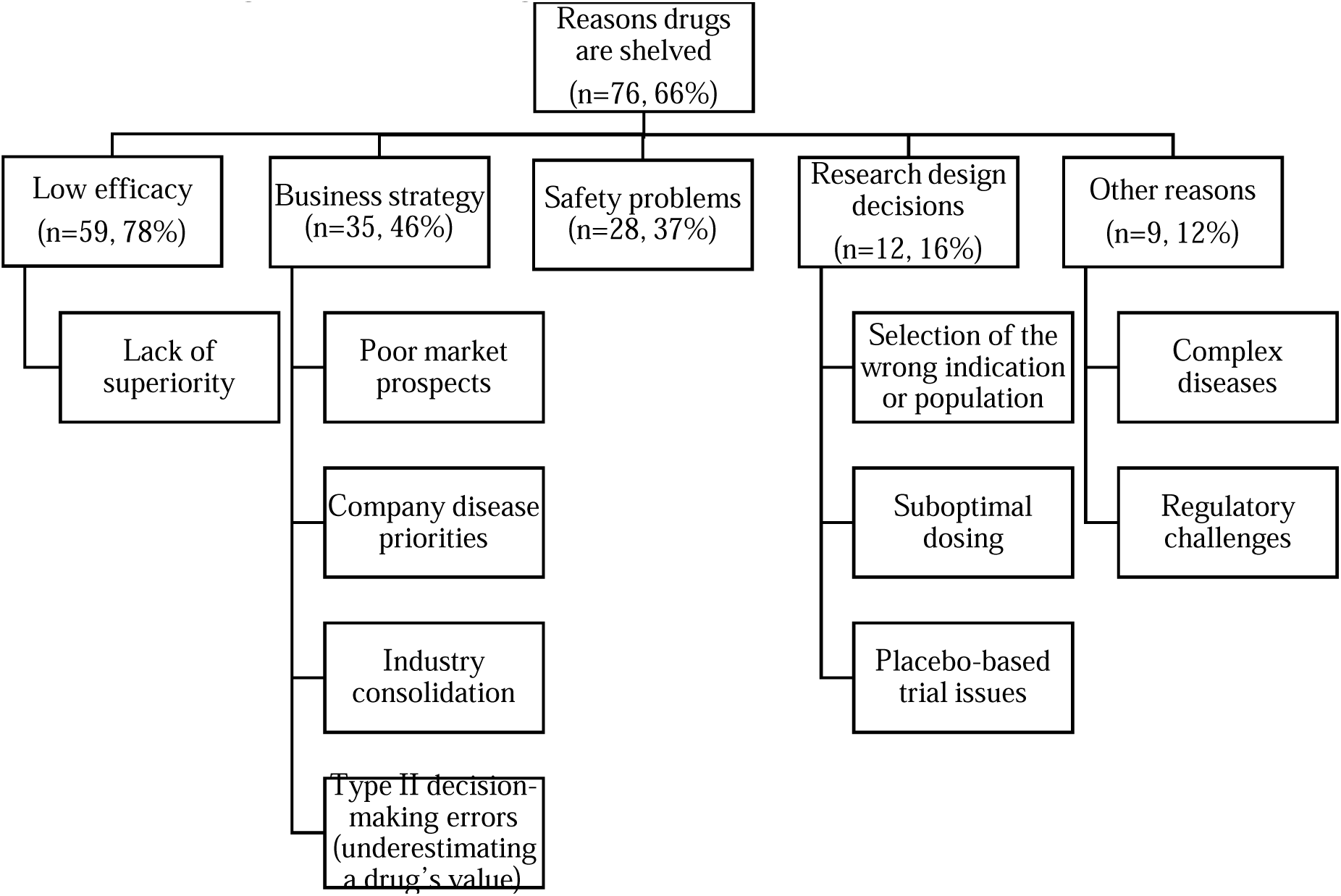
Reasons why a drug’s research and development may be abandoned or shelved

#### Efficacy and superiority challenges

The most frequently cited reason for why drug candidates are abandoned was inadequate efficacy for the studied indication or target population (n=59) or a lack of superiority to alternative therapies (n=11). *Thymitaq*, an experimental cancer drug, for example, was discussed as shelved by Agouron Pharmaceuticals after studies showed it was “clearly active,” but “not sufficiently superior to alternative therapies to justify the required investment [8].” Similarly, imagabalin was discussed as discontinued by Pfizer because it appeared unlikely to “provide meaningful benefit to patients beyond the (then) current standard of care [17].”*Capravirine* was also discontinued after two Phase IIb studies sponsored by Pfizer failed to show a statistically significant difference between standard triple-drug HIV therapies and the same therapy combined with *Capravirine* [9].

#### Strategic business reasons

After efficacy challenges, strategic business reasons were the second most commonly discussed reason for why a promising drug candidate might be abandoned (n=35). Specifically, poor market prospects (n=14), incompatibility with company disease priorities (n=7), industry consolidation (n=5), and type II decision-making errors that can cause a manger(s) to underestimate a drug’s value (n=6) were discussed as leading reasons for a company abandoning development of a promising drug candidate.

Several articles referred to the role of the market in drug development decisions (n=14), though few elaborate on thresholds used to determine the payoff off needed to continue an investment in a drug candidate [10,11,12]. Evaluations of market opportunity can occur at any stage of development, and the risk of commercial embarrassment and financial loss were described as playing large roles in managerial decision making [13]. For example, vaccines were discussed as often abandoned due to a small market and lower revenues, as generally the federal government is the largest purchaser, and frequency use is lower than drugs. Drugs may be used every day, while a vaccine may only be used a few times throughout a person’s lifespan [14].

Further, companies must make prioritization decisions about which compounds to advance given limited resources [13]. If a late-stage compound does not meet set endpoints it is often shelved for the next lead candidate. Re-evaluating a drug’s activity for use in multiple indications is not considered economical [15].

Companies often focus on developing products for specific categories of diseases and conditions. and can abandon drugs targeting conditions outside their research priorities. For example, AstraZeneca sold the rights to its shelved schizophrenia drug candidate to Millendo therapeutics. While the drug was ineffective in schizophrenia, hormonal side effects seen in testing suggested a potential use in polycystic ovary syndrome (POS), which was not a priority therapeutic area for AstraZeneca at the time [16]. Pagoclone (PGC), is another drug that was discussed as abandoned after multiple unsuccessful repurposing efforts by different companies. Studied in 1994 for anxiety by Rhone-Poulenc Rorer, now Sanofi, the drug was later licensed and abandoned by Pfizer due to a lack of robust efficacy data, then pursued by Endo Pharmaceuticals for stuttering and discontinued for reportedly not fitting into the company’s defined R&D priorities and for having low projected commercial potential [17].

Industry consolidation, through for example mergers and acquisitions, can also lead to culling of promising development programs, merging of portfolios, and rivaling factions of scientists [18, 11, 13]. Pfizer, for instance, cut nearly 20% of its development pipeline after acquiring Wyeth in 2009, to help ensure its key disease priority areas were dominant in the new portfolio and to consolidate resources post-merger. This included abandoning imagabalin, under development by Pfizer, reportedly because Pfizer and Wyeth both already had other successful and popular drugs with anxiolytic activity, including Pfizer’s pregabalin and Wyeth’s venlafaxine[17].

After the merger, Pfizer also withdrew its supplemental marketing application for Lyrica to treat anxiety, a drug already approved to treat seizures, fribromyalgia, and nerve pain, among other projects, because they did not fit within their disease and condition priority groups of oncology, pain, inflammation, Alzheimer’s, psychoses, and diabetes. Most abandoned drugs were in phase 1 of development, though three drugs in Phase 2 were also culled. Similarly, Merck cut multiple programs across its pipeline after acquiring Schering-Plough [11].

The literature also noted that managers allocating resources inevitably make errors in their assessment of which projects to continue and which to terminate, especially “type 2 errors” defined as false negative decisions that underestimate a drug’s value; had the organization found the right target and business model, it may have had therapeutic value. These types of judgement errors, where managers underestimate therapeutics’ potential, were discussed as resulting in fewer drugs for patients than would arise in an ideal world [11]. Type 2 errors, that is false negatives, were described as harder to mitigate in comparison to type 1 errors, false positives, which may be caught and or addressed through a rigorous regulatory review. Examples of such errors include the “class effect” (that is negative results for one drug affecting value judgements for others in the same class) and a “felt inferiority” for a compound or “assumption that the compound could be too late to enter the market” [19]. *Dalcetrapib*, for instance, was abandoned by Roche after an independent group stated the drug lacked clinically meaningful efficacy in a late-stage trial. The drug targeted cardiovascular risks, and its failure was discussed as potentially having repercussions for other companies studying similar drugs, like Eli Lilly [20].

#### Research design decisions

Selection of the wrong indication, endpoints, populations to enroll, or patient stratification methodologies in a trial, as well as suboptimal dosing or insensitive biomarkers were discussed as potential drivers of drug abandonment, which we have classified as “research design decisions” (n=12) [21, 22]. One paper, for instance, suggested *Nelivaptan*, a treatment for major depressive disorder and generalized anxiety disorder, may have appeared ineffective because it was studied in the wrong population and for the wrong indication. Study authors noted *Nelivaptan*, an AVPR1b antagonist, would be best utilized in acute stress conditions, as V1b receptors are particularly activated, with limited efficacy for the chronic stress states in which it was tested [21].

In terms of dosing decisions, after initially declaring *Aducanumab* ineffective for treating Alzheimer’s disease after phase III trials, Biogen found statistically significant improvements in cognitive decline in a subset of the sample who had received the highest doses and thus revived the nearly abandoned therapy with new dosage selections [23, 24, 25].There have been over 200 failed Alzheimer’s drugs and candidates, reflecting poorly understood etiology and deficiencies in development and methodology, including issues with dosing, biases, and protocol violations [26]. Papers by Becker described how researchers found several trial related factors in *Phenserine’s* development, also an Alzheimer related drug, that suggested they did not provide fair and unbiased conditions for the drug to demonstrate efficacy, including variance on assessment scores, improvement in the placebo groups, and unaddressed errors [26, 27]. In general, placebo-based trials were discussed as possibly having higher risks for drug abandonment. Comparing a new compound with a placebo was discussed as having a higher risk of a false negative trial, particularly in diseases like irritable bowel syndrome and mild depression, where the placebo responder rate can be as high as 40-50%. Inability to pick a correct dose can also lead to a false-negative effect with placebos, as often the highest acceptable dose, not the most optimal dose, is chosen in order to emphasize the difference versus a placebo [19].

#### Complex Diseases

An inadequate understanding of therapeutic pathways in complex diseases (n=8)- such as Alzheimer’s disease, cardiovascular disease, psychiatric conditions, and stroke- was discussed as a contributor to trial design challenges [22]. In psychiatric disease, for example, inferences from animal research remain limited in scope [28]. In addition, indication selection is relevant as repurposing aims to find new uses for shelved drugs. Importantly, a lack of efficacy for the original indication does not mean a lack of efficacy in other indications. An illustrative example is *Nelivaptan*, which was found to be ineffective as a treatment for major depressive disorder (MDD) and generalized anxiety disorder (GAD). However, nelivaptan is an AVPR1b antagonist and V1b receptors are particularly activated during acute stress, not chronic stress in which it was tested. Thus, *Nelivaptan* is an attractive option for anxiety and disorders of sociality; despite this promising evidence, a company contact suggested that *Nelivaptan* is not available [21]. Lack of efficacy is multifactorial, and interrogating causes of drug abandonment is crucial to demonstrate the potential of repurposing.

Companies were described in the literature as recently drifting away from CNS drug development, as it is now perceived to carry a high risk of failure, despite a high potential reward with a market valued at over $40 billion [29]. High attrition rates in this area reflect issues in translation due to a lack of knowledge of disease etiology and pathology and thus a lack of predictivity of animal models. For example, understanding of the neurophysiology associated with schizophrenia is limited and thus there have been high-profile drug development failures such as the Roche GLYT1 glycine uptake inhibitors. Additionally, despite costly clinical trials of more than 15 neuroprotectant drugs for ischemic stroke, the results were negative [29].

#### Regulatory challenges

Regulatory hurdles (n=2), such as regulators requesting additional studies and a sponsor unwilling to comply, were also discussed as potential drivers for the abandonment of promising drugs [30].

### Barriers to Repurposing

Barriers to repurposing commonly cited in the literature include a lack of finances and resources (n=42), including a lack of expertise (n=11), intellectual property challenges (n=26), poor data access (n=20), bias (n=13) and liability risks (n=5) (See Figure 4). These barriers were discussed as resulting in an unknown number of abandoned compounds stored in company vaults, with some suggesting they may number in the thousands [31, 21].

**Figure 4.**
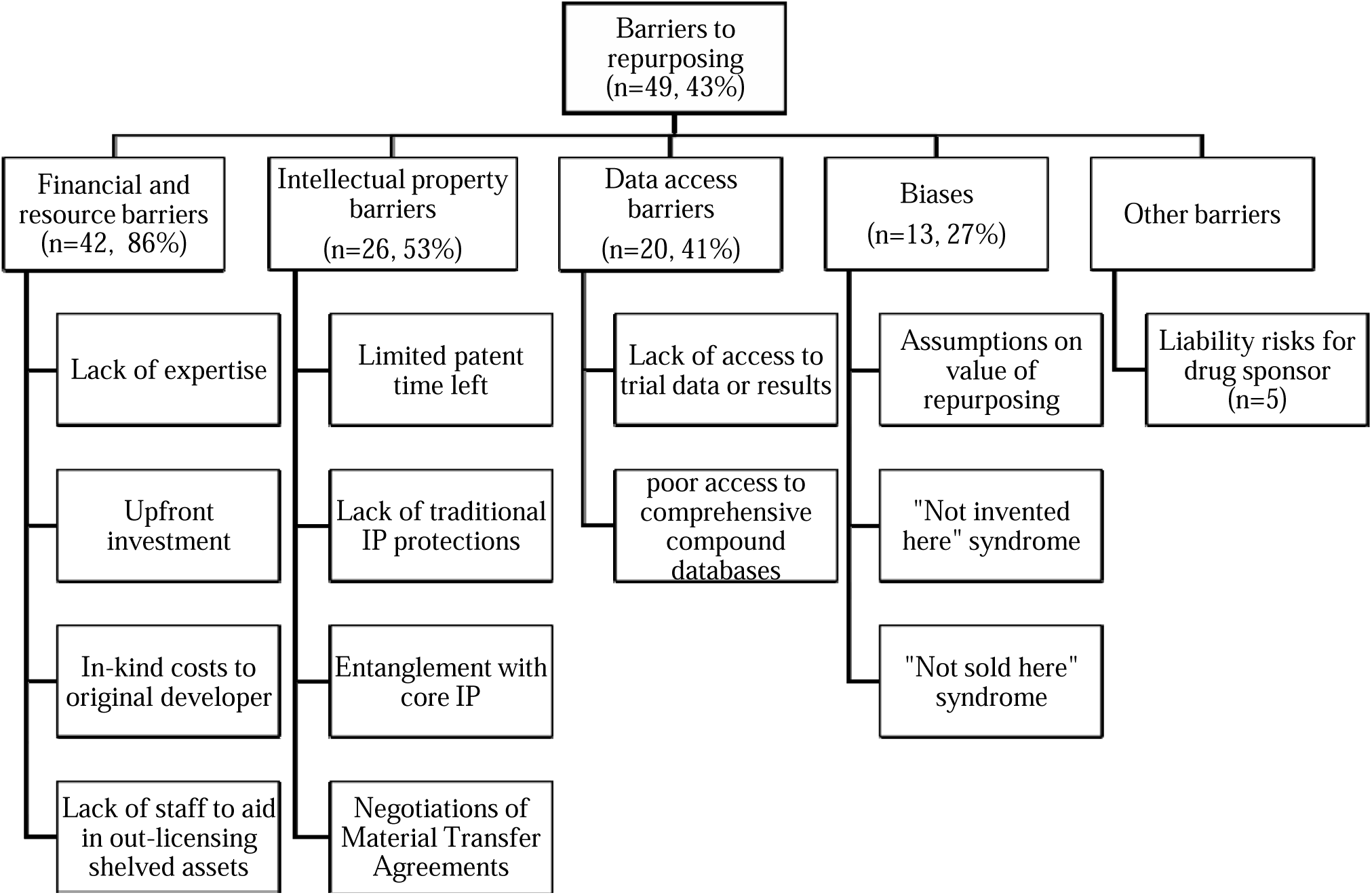
Barriers to repurposing, discussed in the literature

#### Financial and resource barriers

Organizations require financial resources and personnel with relevant expertise on the compound and studied indications to advance a shelved drug candidate. Because pharmaceutical research and development is often organized around specific therapeutic areas within an organization, it can be hard to internally realize the repurposing potential of compounds outside this focus [12, 15]. As such, multi-partner collaborations in repurposing are often needed [32] (Cha, 2018). Academic researchers may have the expertise to study compounds, but not have access to a pool of deprioritized pharmaceutical compounds [33]. Likewise, small biotechnology companies and academic institutions may need to find commercial partners to address a lack of resources [34]. Additionally, companies often lack sufficient staff dedicated to out-licensing discontinued compounds, and thus most are abandoned [35].

Despite the promise of repurposing being a cheaper and faster alternative to de novo development, bringing a repurposed compound to market was described as still costing hundreds of millions to billions of dollars, despite early cost savings from not having to conduct preclinical research [21, 32, 36]. Although many of the compounds have existing data and are well understood, repurposing only reduces, not eliminates drug development risks. Ultimately, repurposing can still require substantive testing, and repurposed compounds must still undergo the same approval process and meet quality, efficacy and safety standards.

Repurposing has been cited as able to save 6-7 years of time spent on preclinical and early-stage research, which may result in millions of dollars’ worth of savings [37]. However, in the later stages, repurposed compounds may still have the same failure rate as any other compound, if not higher, after failing in a primary indication [34, 12]. Repurposed drugs can still require phase 2 and 3 clinical trials, which eliminate 68% and 40% of compounds, respectively, which make it that far, for their new indications [38].

Even when out-licensing a compound, there can be burdensome “in-kind” costs from remanufacturing the active product and placebo, completing study reports and regulatory documentation, pharmacovigilance, monitoring and reporting on patient safety, and coordination [39, 40]. It is challenging to persuade management to allocate resources to compounds that were initially unsuccessful, especially if the new indication is not a strategic focus [12].

#### Intellectual property (IP) barriers

The second most common barrier to drug repurposing discussed in the literature were intellectual property (IP) related. Pharmaceutical companies were described as patenting many compounds, even if they are later abandoned, and thus preventing others from developing these compounds without a license [11]. There can also be limited patent time left for compounds that failed in later stages, limiting return on investments (ROI) in drug repurposing. The threshold companies use to determine if an ROI is worth their investment can vary by company size. What may not be a sufficient ROI for a large company may be enough for a small company and result in a new medication for patients [35].

Further discussed, is a lack of traditional IP protections for repurposed compounds, though products can still be economically successful without this type of protection. Composition of matter (COM) claims are among the most powerful IP protections for newly synthesized compounds. But COM claims can be difficult to gain for repurposed compounds, as the patentee must somehow differentiate their patent claims over what is in the public domain and present data that the drug is a credible candidate for the new indication [41, 42].

Entanglement with core IP is another issue. The literature states it can be common for developers to patent a number of compounds in development, which protects not only the final candidate, but the semi-finalists as well [11]. Thus, shelved compounds from the same family cannot be developed by another party without a license of access to the relevant patents that protect the compounds.

As IP protects pharmaceutical investment and disallows competitors from building upon original research and repurposing compounds, it poses a difficult barrier to address [10, 43]. Material transfer agreements (MTAs) pose a particularly challenging and time-consuming barrier. Negotiations on MTAs are most heated around issues of limiting compound use to non-commercial research, limiting company liability, delaying academic publications to protect confidential information, and IP provisions. IP terms were described as difficult and time-consuming to negotiate as companies often want to protect their freedom to operate using their own compounds, while universities want to maintain ownership of inventions, receive consideration, and make compounds available to the public [44].

#### Data access barriers

Barriers to accessing shelved compounds and their trial data were the second most commonly discussed challenge to drug repurposing. Compounds were described as “disappearing” once their development is abandoned, with trial data and results left unpublished [38, 19]. Several factors were described as influencing trial publication practices around shelved drugs, including the difficulty of publishing negative trial results, that many trials end up terminated abruptly after a company merger or acquisition [17], and the lack of commercial benefit in dedicating time and resources to publishing results on a discontinued project, and no legal requirement to do so [30]. Moreover, data are sometimes sequestered if considered “trade secret” or of potential commercial value.

Gaining knowledge about and access to shelved industry compounds was often described as difficult and, in many cases, requiring an internal company champion for success [40]. Companies were cited as expressing reluctance to share shelved compounds with other companies, in case they turn out to be blockbusters. Nonprofits and government-funded bodies, on the other hand, have a lower risk of commercial embarrassment [13].

Furthermore, a lack of repositories to transparently register abandoned compounds and a reluctance from companies to release compounds to a shared resource were cited as reasons shelved drugs can “disappear”[44]. Within companies, paper records need to be digitized and often the company’s expert on the compound move on and teams responsible for the regulatory and safety data are disbanded [31]. Additionally, mining large datasets poses a logistical hurdle and integration of different types of data in a user friendly manner is challenging [45, 42].

#### Value questions and biases about repurposing

Developers make assumptions on the value of reinvestigating shelved compounds. Some critics have expressed concern that focusing on repurposing detracts from innovation and the pursuit of novel drugs and therefore poses a disservice to the possibility of finding new cures [46]. Some experts also disagree with the notion that shelved drugs represent a significant opportunity for development and report believing there has been “an awful lot of hype” regarding repurposing programs [44, 47]. Addressing value biases were discussed as requiring a great deal of information and is a process that is described as time-consuming and expensive for all involved parties.

The “not sold here” and “not invented here” syndromes were discussed. The “not sold here” syndrome refers to the unwillingness of companies to out-license compounds that may be promising for other indications [11]. Business units argue that if they do not sell a product, no one else should, leading to a waste of human talent in research and development as compound attrition rates are quite high. The “not invented here” syndrome refers to the bias that external research and technology are inferior to a company’s own R&D capabilities and standards and therefore not worth pursuing. External parties may also assume a seller is keeping the best compounds for themselves and offering lesser value compounds for out-licensing [11].

Pharmaceutical companies in general were described as employing few, if any, staff to aid in out-licensing shelved compounds which limits outside companies’ evaluation of shelved drugs.

#### Liability risks

Drug companies may also face liability risks (n=5) when out-licensing abandoned compounds, which include risks of adverse patient events, a need to continuously supply the compound to the licensee, and litigation [11]. Testing compounds for new indications and populations may reveal new adverse events or unforeseen toxicities [48]. For externally sponsored studies, the investigator must report back safety data to the parent company.

The high cost of liability insurance was discussed as a reason pharmaceutical companies discontinue development of lower-revenue products like vaccines. To meet the demand for increased liability insurance, the cost of the pertussis vaccine rose from 17 cents to 11 dollars per dose, and the number of companies making the vaccine reportedly decreased [14].

#### Facilitators of Repurposing

The most common facilitators of drug repurposing discussed were collaborative initiatives (n=38), compound libraries and databases (n=24), computational based strategies and tools (n=32), regulatory modifications (n=5), and tax incentives (n=2) (See Figure 5).

**Figure 5.**
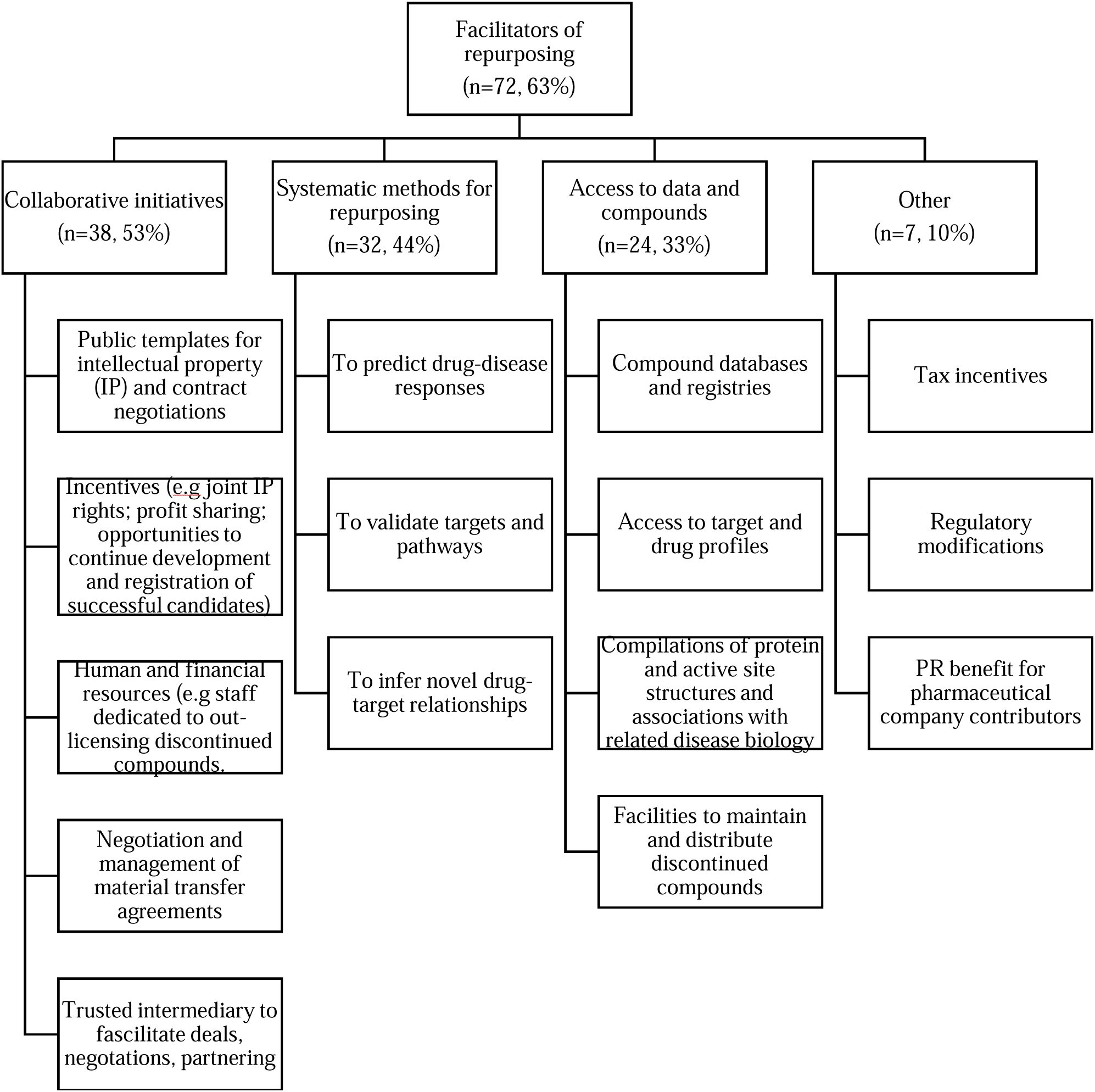
Facilitators of drug repurposing, discussed in the literature

#### Collaborative initiatives

Multi-partner collaborations between pharmaceutical companies and academic institutions, non-profit organizations and biotechnology companies was the most commonly discussed facilitator for drug repurposing cited in the literature (n=38). Pharmaceutical companies are often perceived as having the resources as well as a pool of shelved compounds and data while biotechnology companies and academia as having knowledge and expertise on emerging areas to study compounds and contribute to innovation [49]. A staff dedicated to out-licensing discontinued compounds was described as a facilitator for collaborative partnerships [35].

Several examples of collaborative initiatives focused on drug repurposing were discussed in the literature, including the NIH National Center for Advancing Translational Sciences (NCATS) program: Discovering New Therapeutic Uses for Existing Molecules (n=23), the Medical Research Council (MRC) and AstraZeneca (AZ) Mechanisms of Human Disease Initiative, (n=16), the AZ Open Innovation program (n=6), European College of Neuropsychopharmacology (ECNP) Medicines Chest Program (n=5), The Clinical and Translational Science Award (CTSA) Pharmaceutical Assets Portal (2 n=), Pfizer’s SpringWorks Program (n=2), the AstraZeneca/National Research Program for Biopharmaceuticals(NRPB) partnership in Taiwan (n=2), the Clinical Development Partnerships Initiative (n=1), the Roche/Broad Institute Collaboration (n=1), and the Drugs for Neglected Diseases Initiative (DNDI) (n=1).

The NIH NCATS’ Discovering New Therapeutic Uses for Existing Molecules program was initiated in 2012 to help scientists explore new treatments for patients by matching NIH-funded researchers with a selection of 58 compounds previously discontinued from development due to lack of efficacy, selectivity, or strategy [36, 39, 50]. Co-launched with AstraZeneca (AZ), Eli Lilly, and Pfizer, the initiative required that compounds had prior evidence and manageable tolerability in humans and that companies publicly posted online template confidentiality disclosure agreement (CDAs) and collaborative research agreements (CRAs) to enable rapid implementation [39, 51]. In the program, the NIH acts as a trusted intermediary, facilitating deals between researchers and companies that are often characterized by prolonged negotiation, and moving promising compounds into the private sector [36, 42, 52, 53]. Organizations maintain IP on the original compound, but the repurposed use belongs to the researchers. However, companies can license the IP from researchers, and researchers can request licenses for additional studies [52]. In short, the NIH NCATS initiative facilitates the availability of compounds, data, human, and financial resources, and addresses issues of intellectual property and data sharing in drug repurposing [31].

Another partnership between the MRC and AZ, the Mechanisms of Human Disease Initiative, launched in 2011 provided researchers with what was described as “unprecedented access” to clinical and pre-clinical AstraZeneca compounds. It accepted proposals for novel research projects with a focus on understanding human disease [38]. The MRC posted data on 22 compounds on its website, and over 100 proposals were submitted from across the UK. Full proposals were developed by UK researchers and AZ scientists, and selected proposals were funded by the MRC [39]. AstraZeneca also launched another program with the National Research Program for Biopharmaceuticals (NRPB) in Taiwan to facilitate translational research locally, which included live compounds [42]. As a result of the success of pilot programs, AstraZeneca launched an Open Innovation program that seeks to make a range of unwanted molecules readily available to university researchers who can propose novel repositioning ideas (Frail). In doing so, AstraZeneca gains a competitive advantage when the same scientists are looking for companies to share breakthroughs with [16].

The European College of Neuropsychopharmacology (ECNP) Medicines Chest Program was set up to provide academic and small company researchers access to promising compounds for experimental medicine studies. Similar to the NIH NCATS program, the compounds are placed on the ECNP website, and researchers are invited to submit a 2-3 page proposal outlining aclinical study. After the ECNP vets the study, a contract, of which a sample is publicly available, is drawn up between the company and academic institutions and access to confidential information is provided for grant applications to fund the study [40].

The Clinical and Translational Science Award (CTSA) Pharmaceutical Assets Portal facilitates industry-academic collaborations for discovery of new indications for shelved compounds by providing a foci-of-expertise tool that identifies investigators with complementary interests, access to resource-management tools, facilities to house, maintain and distribute the discontinued compounds, management of IP and material transfer agreements, and selection of projects for funding [44, 52].

Likewise, The Clinical Development Partnerships Initiative presents a cost-effective, rapid means by which pharmaceutical companies can boost their product lines. Companies retain IP rights to their original molecule and first option to view trial data if they loan their compounds to Cancer Research UK, which will conduct early phase I and II clinical trials. The company retains the option to develop and market the drug, and the charity receives a share of any revenue [13].

The Roche/Broad Institute Collaboration made 300 compounds which failed to meet critical phase II milestones or were shelved for strategic reasons available to researchers who could suggest experiments. If collaborators uncovered any shared findings, Roche and the partner would agree on next steps, including publishing results, further experimentation, or a development plan [52].

Lastly, the Drugs for Neglected Diseases Initiative (DNDI) is an open-source collaborative endeavor that partners the expertise and assets of pharmaceutical companies with networks of public and private scientists to support repurposing and investigation of novel treatments for neglected tropical diseases. Merck entered a collaboration with DNDI in which it would provide small molecule assets and respective intellectual property through socially responsible licensing agreements to develop, manufacture and distribute cost-effective treatments for NTDs to resource-poor countries. The pharma company would share joint IP rights on candidates in early development, with an opportunity to continue late clinical development and registration of successful candidate. In doing so, collaboration is incentivized and resident expertise and contributions in later stages of development help maximize the drug’s potential [54].

#### Databases and registries

Compound access is another important facilitator for repurposing. Many initiatives serve to create databases which provide target and drug profiles, including protein and active site structures and associations with related diseases and biological functions, to interested researchers. Databases discussed in the literature include PubChem (7), DrugBank (6), Promiscuous (5), ChEMBL(5), the NIH clinical collection (2), the Open Phacts Initiative (2), DisGenNet (1), the Drug Repurposing Hub (1), DrugSig (1), and the US FDA’s Orange Book of discontinued drug products list (1).

Compound-specific databases include: PubChem, which is administered by the NIH, holds data from several hundred biochemical and phenotypic screens, with more deposited each month [55], ChEMBL, an open target platform that enables investigation of evidence-associated targets and disease in an accessible manner by presenting molecules with drug-like properties [56] and the US FDA’s Orange Book of discontinued drug products.

DrugBank is the most comprehensive publicly available database of approved, experimental and withdrawn drugs which are annotated by indication and intended targets [57]. The Promiscuous database provides an exhaustive set of drugs (25,000), including withdrawn or experimental drugs with drug-protein and protein-protein relationships annotated, allowing researchers to identify prospective new uses by examining predictive interaction points [52]. The NIH clinical collection presents a library of drugs that passed safety tests but for various reasons did not reach the market [38]. The Open Phacts initiative allows for multiple sources of publicly available pharmacological and physicochemical data to be intuitively queried, with 28 partners from public and private sectors [52]. DisGeNet offers associations between genes and diseases, as well as disease-variant associations.

The Drug Repurposing Hub is a database of approved, clinical-trial tested, and pre-clinical compounds that are annotated with literature-reported targets, and DrugSig is a public resource for signature-based drug repositioning that builds off of the Connectivity Map from the Broad Institute [57]. Open-source databases allow for efficient sharing of resources, compounds and drugs to cost-effectively advance shelved compounds, and provide a PR benefit for pharmaceutical contributors, who are not locked into long-term commitments.

#### Systematic methods for repurposing

Many novel methods have been developed and applied to help identify and validate repurposing targets, greatly advancing repurposing endeavors (n=32). Computational approaches coupled with open-access databases were described as central in identifying potential repurposing opportunities by predicting drug-disease responses and validating targets and pathways [21].

Of these newer methods, signature-based approaches (n=5) were commonly employed for drug repurposing. These include investigating published GWAS data from institutes like the US National Human Genome institute to systematically and rapidly identify alternative indications for existing drugs and exploring how many genes are amenable to pharmacological intervention using biopharmaceuticals or small molecules [58]. However, key limiting factors are the expertise and time required to develop such assays and integrating databases that identify known drugs among confirmed activities [55].

In-silico screening of compound libraries (n=4) is useful in both significantly reducing wet-laboratory work and lowering the cost of experimental determination of drug-target interactions [59]. Additionally, public access to high throughput screens (HTS) of small molecules (n=3), particularly mining of phenotypic screens, was described as an effective and economical strategy for repurposing drugs [55].

Computer-aided approximations include: bioinformatics-based approaches (n=3) which employ domain similarity prediction tools and sequence alignment to discover novel protein-protein similarities, identifying closely related targets and new repurposing opportunities, and chemoinformatics-based approaches (n=2) which involve molecular representations of candidate compounds which are submitted to computational algorithms which rank and prioritize compounds for experimental testing, When 3D structures are available, molecular docking (n=1) can be used to screen a large number of compounds against a target protein. When they are not, ligand and network-based approaches can be utilized [59].

Network modeling (n=1) and systems-biology approaches (n=1), were also discussed as helpful. Network modeling reconstructs a biological network and simulates its interactions to reveal potential drug targets [60, 61]. A systems-biology approach was described as the use of omics data, signaling pathways, metabolic pathways and protein interactions to come up with a new pathway for a proposed disease [43].

While most network-based approaches are limited in their predictions of how drugs and targets interact, machine learning approaches (n=3) can go further in accurately predicting drug-target interactions and inferring modes of action and novel drug-target relationships [59].

Finally, AI-driven technology (n=1) can integrate diverse types of data, and look for connections. For example, Biovista has developed an AI solution called Project Prodigy which does not limit itself to machine learning but rather is capable of building entirely new clinical scenarios and has led to internal repurposing successes in multiple sclerosis and epilepsy. Their AI system has been used in collaboration with major pharmaceutical companies, patient advocacy groups, and the FDA [62].

#### Tax incentives and regulatory modifications

Tax incentives and certain regulatory modifications may further facilitate drug repurposing. Tax incentives such as allowing for the deduction of residual product value upon donation of a compound or for sharing trial data to a third party could help advance development of shelved compounds. Regulatory modifications to the FDA’s 505(b)(2) pathway that allow for use of previously compiled, but not previously FDA evaluated, safety data. Use of the FDA’s safety findings could expand the number of drugs available without adversely impacting risk benefit [21].

### Examples of successfully repurposed or re-positioned drugs

The most frequently discussed repurposing opportunities were for rare and neglected diseases (n=12), Alzheimer’s disease (n=10), AIDS (n=2), and central nervous system disorders (n=2). Examples of successfully repositioned drugs (n=50) discussed in the literature included thalidomide (n=8), Viagra/slidenafil (n=7), Saractinib (6), AZT (n=5), Aducanumab (n=4), Sunitinib (n=3), Ebselen (n=2), tamoxifen (n=2), raloxifene (n=2) and daptomycin (n=1).

Drug promiscuity, the notion that one drug can affect more than a single pathway and lead to new indications for drug candidates was frequently discussed, with thalidomide the most commonly given example [61]. Thalidomide (n=8), originally manufactured by the German company Chemie Grunenthal in the mid 1950’s, was discussed as an example of a drug that failed after its market launch in several countries, though it was not approved by the US FDA at the time, and later successfully repurposed [11]. Originally indicated for sedation and morning sickness, it was withdrawn for its teratogenic effects. However, further studies revealed that the drug inhibited tumor necrosis factor-alpha signaling, and was subsequently approved for the treatment of erythema nodosum leprosum, a life threatening complication of leprosy, and then multiple myeloma. In the US, the FDA approved the drug for acute ENL, in 1998, however, “use was limited by very strict guidelines,” according to the literature [11].

Viagra was also presented in the literature as a well-known example of a drug that showed lack of efficacy in clinical trials for its originally studied indication, but interestingly, analysis of its unusual side effects and its poor pharmacokinetic properties for angina led to its eventual use for erectile disfunction [11, 61].

Daptomycin, an antibiotic, was successfully repurposed by Cubist after Eli Lilly abandoned it when downsizing its infectious disease division. Eli Lilly out licensed the drug to Cubist after four years on the shelf. Cubist’s Chief scientific officer advocated for use of daptomycin as an antibiotic. IP negotiations proved challenging, but eventually Cubist purchased worldwide development and commercialization rights to Daptomycin along with a license to the underlying IP related to the compound, and Eli Lilly has received over 333$ million in royalties on the product sales to date [11]. Cubist is described as redesigned the clinical trials and filing a patent on the basis of a once-daily treatment regimen to minimize adverse effects from the drug. Daptomycin is now an important public health tool, serving as a last resort medication proven useful in diseases like MRSA that have become increasingly resistant to front line antibiotics.

Saracitinib was originally developed for multiple oncology indications, but phase II studies showed limited benefit and the drug was deprioritized. The concept for repositioning of this agent came from discoveries of memory impairments in mouse models of AD and data that showed the phosphorylation of the Fyn tyrosine kinase was related to Aβ- and tau-associated synaptic dysfunction. The drug is currently being investigated for other indications like bone pain and lymphangioleiomyomatosis through MRC, NIH, and NCATS programs [39, 63, 64].

Azidothymidine (AZT) likewise reflects how a detailed understanding of disease and drug mechanisms of action can lead to repurposing discoveries in entirely new indications. AZT was originally investigated as a chemotherapy drug in the 1960’s but was abandoned due to lack of efficacy. However, in the early days of the HIV epidemic, AZT’s anti-retroviral effect was noted, and the NIH partnered with industry experts to repurpose the drug, which became the first treatment for patients with HIV [32, 38, 65, 66].

Aducanumab was abandoned after a futility analysis from an independent monitoring committee indicated the drug was not going to be effective for treating Alzheimer’s disease. However, a re-analysis of data from two failed clinical trials showed promising results, as a subset of patients treated with the highest dose appeared to show a statistically significant slowing of decline of cognitive ability and basic activities of daily living. Biogen concluded the initial analysis had been incorrect and got support from the FDA to move forward with a regulatory filing, reviving the nearly abandoned drug [23, 24, 25, 67].

Sunitinib presents an example of on-target repurposing. It failed in clinical trials for colorectal, breast, prostate, and non-small cell lung cancer, but was successfully repositioned for treatment of gastrointestinal stromal tumor and renal cancers, and after a repurposing effort approved for treatment of pancreatic neuroendocrine tumors in 2010 [61]. Analysis of the lack of efficacy of Sunitinib in some cancers demonstrates the importance of a targeted approach [68].

A drug repurposing approach screening of the National Health Institute Clinical Collection identified Ebselen as a potential lithium mimetic [69]. Ebselen was originally indicated for stroke, but showed a lack of efficacy. Never marketed, Ebselen could have repurposing potential for treatment of bipolar disorder, and in a paper published in 2016 was described as currently under investigation [38].

Tamoxifen was a failed contraceptive and orphan drug, though in translational laboratory work it showed efficacy in induction of ovulation in sub-fertile women and in the treatment of metastatic breast cancer in postmenopausal women. A nonsteroidal antiestrogen, tamoxifen was repurposed and approved for treatment of metastatic breast cancer and later for breast cancer risk reduction, and is currently the standard of care for long term adjuvant therapy for estrogen receptor positive breast cancer [70]. A cluster of translational studies around the 1970’s and 80’s focused on the uterus, breast and bone created a database for further studies and trials that also resulted in the reinvention of keoxifene, a failed breast cancer drug, to raloxifene, the first clinically available selective estrogen receptor modulator for breast cancer and osteoporosis prevention [70].

## DISCUSSION

In this systematic literature review, we examined why pharmaceutical companies de-prioritize, shelve or abandon development of promising drug candidates as well as facilitators and barriers for successful repurposing of promising compounds.

We found the most commonly discussed reasons for why a promising drug may be abandoned were inadequate efficacy, or superiority over other therapies, for the studied indication or population, followed by strategic business reasons by the sponsor often related to judgments about a drug’s market prospects or industry consolidation, as well as flawed research design decisions. Inadequate understanding of therapeutic pathways in complex diseases, such as for Alzheimer’s disease, cardiovascular disease, psychiatric conditions, and stroke were presented as compounding trial design challenges. In psychiatric disease, for example, inferences from animal research were discussed as remaining limited in scope. Regulators requesting the completion of additional studies and a sponsor unwilling to comply, were also discussed as potential drivers for drug abandonment. These findings support a previous study evaluating why clinical stage compounds that have cleared regulatory review in Phase 1 safety trials are subsequently abandoned before reaching the market, which found 38% were due to inadequate efficacy for the studied disease, 34% due to poor perceived economics, 20% for safety reasons, and 9% for other reasons.

The top barrier to drug repurposing was inadequate resources, especially financial, subject matter expertise, and dedicated staff focused on out-licensing. IP challenges and inadequate data access were among other leading barriers as well as value questions and assumptions on the role of repurposing as an effective tool in drug development. While some papers describe drug repurposing as faster, cheaper, and more likely to succeed than traditional drug development approaches, others note that in later stages, repurposed compounds may still have the same failure rate as any other compound, if not higher, after failing in a primary indication. Liability risks were also presented as barriers to re-purposing. Altogether, these barriers were presented as resulting in an unknown number of abandoned compounds stored in company vaults, with some suggesting they may number in the thousands [21, 31].

The most common facilitators for repurposing, we found, were collaborative partnerships between bio-pharmaceutical companies, academia, and non-profit organizations that help bring together needed resources and expertise. Of note, the role of patients and patient organizations as collaborators in drug repurposing was largely unaddressed in reviewed literature, despite their growing role in more traditional forms of drug research and development [123]. Access to compound libraries and databases, the development and application of new computational methods to screen databases, regulatory modifications, and tax incentives were also identified as facilitators. Many of these facilitators generally correlate, as opposites, to the barriers we found to re-purposing.

However, a major barrier also includes successful negotiation of material transfer agreements between potential collaborators, an issue without a clear solution in the literature, suggesting a need for further study on ways to better support IP negotiations to more fully realize repurposing benefits. There are some models in the literature, such as the NIH NCATS repurposing program, which may be offer helpful generalizable best practices for supporting IP negotiations. The program was described as allowing the NIH to act as a trusted intermediary with procedures for facilitating deals, including on IP, between researchers and companies.

Further, the literature emphasizes biases around the value of re-purposing in drug development, as a barrier to re-purposing. We found these value questions reflected in our findings, as some papers described repurposing as faster, cheaper, and more likely to succeed than traditional drug development approaches, while others argue in later stages, repurposed compounds may still have the same failure rate as any other compound, if not higher, after failing in a primary indication. More systematic study may be needed on the current value of repurposing as a tool in drug development.

There are limitations to this study. Notably, included publications were often descriptive papers and perspective pieces, not rigorous scientific studies, which limits the conclusions that can be drawn. This review excluded one paper that was not available in English and did not search any non-English database. Conference abstracts were excluded due to insufficient extractable data. Additionally, information on the characteristics we were abstracting about drug abandonment and repurposing may not have been published in the medical or pharmaceutical peer-review literature and may have been missed.

## CONCLUSION

In this systematic literature review assessing why development of promising drug candidates are abandoned, we found insufficient efficacy, or superiority to other therapies, for studied indications or populations, judgements about a product’s market prospects and industry consolidation among leading factors. Inadequate resources and challenges negotiating IP and data access are key barriers needing reform for repurposing to reach its full potential as a core approach in drug development. Multi-partner collaborations, along with the creation, accessibility, and use of compound databases, regulatory modifications and tax incentives are key facilitators for repurposing promising shelved drugs. More research is needed on the current value of repurposing as a core method in drug development and how to better facilitate resources to support it, where valuable, especially financial, staffing focused on out-licensing shelved products, and legal expertise to negotiate IP agreements in multi-partner collaborations.

## Data Availability

The analyzable dataset will be shared on dryad.

## Declarations

An ethics review was not needed for this research, as no human subjects were involved. The analyzable dataset will be shared on dryad. The authors have the following competing interests to disclose, Dr. Miller receives grant funding from the National Institutes of Health, Susan G. Komen Foundation, Milken Institute, and Arnold Ventures, serves on the Alexion bioethics advisory committee, board of the nonprofit Bioethics International, and Cambia Health pharmacy and therapeutics committee. This research study was funded by a grant from Faster Cures of the Milken Institute. NK, AG, SA and JEM designed this study as well as extracted and analyzed data. All authors contributed to the drafting, revising and approving of this paper for publication.

