## Appendix for "Why promising drugs are shelved and barriers and facilitators to re-purposing them: A systematic literature review"

### Supplementary Table 1: PRISMA 2020 Main Checklist

| **Topic** | **No.** | **Item** | **Location where item is reported** |
| --- | --- | --- | --- |
| **TITLE** |  |  |  |
| **Title** | 1 | Identify the report as a systematic review. | LN1-2 |
| **ABSTRACT** |  |  |  |
| **Abstract** | 2 | See the PRISMA 2020 for Abstracts checklist | Supp. |
| **INTRODUCTION** |  |  |  |
| **Rationale** | 3 | Describe the rationale for the review in the context of existing knowledge. | LN21-45 |
| **Objectives** | 4 | Provide an explicit statement of the objective(s) or question(s) the review addresses. | LN47-53 |
| **METHODS** |  |  |  |
| **Eligibility criteria** | 5 | Specify the inclusion and exclusion criteria for the review and how studies were grouped for the syntheses. | LN77-84 |
| **Information sources** | 6 | Specify all databases, registers, websites, organisations, reference lists and other sources searched or consulted to identify studies. Specify the date when each source was last searched or consulted. | LN61-69 |
| **Search strategy** | 7 | Present the full search strategies for all databases, registers and websites, including any filters and limits used. | Supp. 2 |
| **Selection process** | 8 | Specify the methods used to decide whether a study met the inclusion criteria of the review, including how many reviewers screened each record and each report retrieved, whether they worked independently, and if applicable, details of automation tools used in the process. | LN71-77 |
| **Data collection process** | 9 | Specify the methods used to collect data from reports, including how many reviewers collected data from each report, whether they worked independently, any processes for obtaining or confirming data from study investigators, and if applicable, details of automation tools used in the process. | LN87-89 |
| **Data items** | 10a | List and define all outcomes for which data were sought. Specify whether all results that were compatible with each outcome domain in each study were sought (e.g. for all measures, time points, analyses), and if not, the methods used to decide which results to collect. | LN89-94 |
|  | 10b | List and define all other variables for which data were sought (e.g. participant and intervention characteristics, funding sources). Describe any assumptions made about any missing or unclear information. | LN94-96 |
| **Study risk of bias assessment** | 11 | Specify the methods used to assess risk of bias in the included studies, including details of the tool(s) used, how many reviewers assessed each study and whether they worked independently, and if applicable, details of automation tools used in the process. | LN94-96 |
| **Effect measures** | 12 | Specify for each outcome the effect measure(s) (e.g. risk ratio, mean difference) used in the synthesis or presentation of results. | N/A |
| **Synthesis methods** | 13a | Describe the processes used to decide which studies were eligible for each synthesis (e.g. tabulating the study intervention characteristics and comparing against the planned groups for each synthesis (item 5)). | LN89-94 |
|  | 13b | Describe any methods required to prepare the data for presentation or synthesis, such as handling of missing summary statistics, or data conversions. | LN89-94 |
|  | 13c | Describe any methods used to tabulate or visually display results of individual studies and syntheses. | LN89-94 |
|  | 13d | Describe any methods used to synthesize results and provide a rationale for the choice(s). If meta-analysis was performed, describe the model(s), method(s) to identify the presence and extent of statistical heterogeneity, and software package(s) used. | N/A |
|  | 13e | Describe any methods used to explore possible causes of heterogeneity among study results (e.g. subgroup analysis, meta-regression). | N/A |
|  | 13f | Describe any sensitivity analyses conducted to assess robustness of the synthesized results. | N/A |
| **Reporting bias assessment** | 14 | Describe any methods used to assess risk of bias due to missing results in a synthesis (arising from reporting biases). | N/A |
| **Certainty assessment** | 15 | Describe any methods used to assess certainty (or confidence) in the body of evidence for an outcome. | LN94-96 |
| **RESULTS** |  |  |  |
| **Study selection** | 16a | Describe the results of the search and selection process, from the number of records identified in the search to the number of studies included in the review, ideally using a flow diagram. | LN100-105 |
|  | 16b | Cite studies that might appear to meet the inclusion criteria, but which were excluded, and explain why they were excluded. | Suppl. 3 |
| **Study characteristics** | 17 | Cite each included study and present its characteristics. | Suppl. 4 |
| **Risk of bias in studies** | 18 | Present assessments of risk of bias for each included study. | Suppl. 4 |
| **Results of individual studies** | 19 | For all outcomes, present, for each study: (a) summary statistics for each group (where appropriate) and (b) an effect estimate and its precision (e.g. confidence/credible interval), ideally using structured tables or plots. | LN108-676 |
| **Results of syntheses** | 20a | For each synthesis, briefly summarise the characteristics and risk of bias among contributing studies. | Suppl. 4 |
|  | 20b | Present results of all statistical syntheses conducted. If meta-analysis was done, present for each the summary estimate and its precision (e.g. confidence/credible interval) and measures of statistical heterogeneity. If comparing groups, describe the direction of the effect. | N/A |
|  | 20c | Present results of all investigations of possible causes of heterogeneity among study results. | N/A |
|  | 20d | Present results of all sensitivity analyses conducted to assess the robustness of the synthesized results. | N/A |
| **Reporting biases** | 21 | Present assessments of risk of bias due to missing results (arising from reporting biases) for each synthesis assessed. | N/A |
| **Certainty of evidence** | 22 | Present assessments of certainty (or confidence) in the body of evidence for each outcome assessed. | N/A |
| **DISCUSSION** |  |  |  |
| **Discussion** | 23a | Provide a general interpretation of the results in the context of other evidence. | LN679-698 |
|  | 23b | Discuss any limitations of the evidence included in the review. | LN714-716 |
|  | 23c | Discuss any limitations of the review processes used. | LN716-719 |
|  | 23d | Discuss implications of the results for practice, policy, and future research. | LN722-730 |
| **OTHER INFORMATION** |  |  |  |
| **Registration and protocol** | 24a | Provide registration information for the review, including register name and registration number, or state that the review was not registered. | LN57-58 |
|  | 24b | Indicate where the review protocol can be accessed, or state that a protocol was not prepared. | LN17 |
|  | 24c | Describe and explain any amendments to information provided at registration or in the protocol. | N/A |
| **Support** | 25 | Describe sources of financial or non-financial support for the review, and the role of the funders or sponsors in the review. | LN |
| **Competing interests** | 26 | Declare any competing interests of review authors. | LN |
| **Availability of data, code and other materials** | 27 | Report which of the following are publicly available and where they can be found: template data collection forms; data extracted from included studies; data used for all analyses; analytic code; any other materials used in the review. | LN57-58 |

*From:* Page MJ, McKenzie JE, Bossuyt PM, Boutron I, Hoffmann TC, Mulrow CD, et al. The PRISMA 2020 statement: an updated guideline for reporting systematic reviews. MetaArXiv. 2020, September 14. DOI: 10.31222/osf.io/v7gm2. For more information, visit: www.prisma-statement.org

### PRIMSA Abstract Checklist

| **Topic** | **No.** | **Item** | **Reported?** |
| --- | --- | --- | --- |
| **TITLE** |  |  |  |
| **Title** | 1 | Identify the report as a systematic review. | Yes |
| **BACKGROUND** |  |  |  |
| **Objectives** | 2 | Provide an explicit statement of the main objective(s) or question(s) the review addresses. | Yes |
| **METHODS** |  |  |  |
| **Eligibility criteria** | 3 | Specify the inclusion and exclusion criteria for the review. | Yes |
| **Information sources** | 4 | Specify the information sources (e.g. databases, registers) used to identify studies and the date when each was last searched. | Yes |
| **Risk of bias** | 5 | Specify the methods used to assess risk of bias in the included studies. | Yes |
| **Synthesis of results** | 6 | Specify the methods used to present and synthesize results. | Yes |
| **RESULTS** |  |  |  |
| **Included studies** | 7 | Give the total number of included studies and participants and summarise relevant characteristics of studies. | Yes |
| **Synthesis of results** | 8 | Present results for main outcomes, preferably indicating the number of included studies and participants for each. If meta-analysis was done, report the summary estimate and confidence/credible interval. If comparing groups, indicate the direction of the effect (i.e. which group is favoured). | Yes |
| **DISCUSSION** |  |  |  |
| **Limitations of evidence** | 9 | Provide a brief summary of the limitations of the evidence included in the review (e.g. study risk of bias, inconsistency and imprecision). | Yes |
| **Interpretation** | 10 | Provide a general interpretation of the results and important implications. | Yes |
| **OTHER** |  |  |  |
| **Funding** | 11 | Specify the primary source of funding for the review. | Yes |
| **Registration** | 12 | Provide the register name and registration number. | Yes |

**Synthesis Without Meta-analysis (SWiM) reporting items**

The citation for the Synthesis Without Meta-analysis explanation and elaboration article is: Campbell M, McKenzie JE, Sowden A, Katikireddi SV, Brennan SE, Ellis S, Hartmann-Boyce J, Ryan R, Shepperd S, Thomas J, Welch V, Thomson H. Synthesis without meta-analysis (SWiM) in systematic reviews: reporting guideline BMJ 2020;368:l6890 <http://dx.doi.org/10.1136/bmj.l6890>

| **SWiM is intended to complement and be used as an extension to PRISMA** | | | |
| --- | --- | --- | --- |
| **SWiM reporting item** | **Item description** | **Page in manuscript where item is reported** | **Other*** |
| *Methods* | | | |
| **1** Grouping studies for synthesis | 1a) Provide a description of, and rationale for, the groups used in the synthesis (e.g., groupings of populations, interventions, outcomes, study design) | 3 |  |
|  | 1b) Detail and provide rationale for any changes made subsequent to the protocol in the groups used in the synthesis | N/A |  |
| **2** Describe the standardised metric and transformation methods used | Describe the standardised metric for each outcome. Explain why the metric(s) was chosen, and describe any methods used to transform the intervention effects, as reported in the study, to the standardised metric, citing any methodological guidance consulted | N/A |  |
| **3** Describe the synthesis methods | Describe and justify the methods used to synthesise the effects for each outcome when it was not possible to undertake a meta-analysis of effect estimates | 3 |  |
| **4** Criteria used to prioritise results for summary and synthesis | Where applicable, provide the criteria used, with supporting justification, to select the particular studies, or a particular study, for the main synthesis or to draw conclusions from the synthesis (e.g., based on study design, risk of bias assessments, directness in relation to the review question) | 3 |  |

| **SWiM reporting item** | **Item description** | **Page in manuscript where item is reported** | **Other*** |
| --- | --- | --- | --- |
| **5** Investigation of heterogeneity in reported effects | State the method(s) used to examine heterogeneity in reported effects when it was not possible to undertake a meta-analysis of effect estimates and its extensions to investigate heterogeneity | 3 |  |
| **6** Certainty of evidence | Describe the methods used to assess certainty of the synthesis findings | 3 |  |
| **7** Data presentation methods | Describe the graphical and tabular methods used to present the effects (e.g., tables, forest plots, harvest plots).  Specify key study characteristics (e.g., study design, risk of bias) used to order the studies, in the text and any tables or graphs, clearly referencing the studies included | 3 |  |
| *Results* | | | |
| **8** Reporting results | For each comparison and outcome, provide a description of the synthesised findings, and the certainty of the findings. Describe the result in language that is consistent with the question the synthesis addresses, and indicate which studies contribute to the synthesis | 3-18 |  |
| *Discussion* |  |  |  |
| **9** Limitations of the synthesis | Report the limitations of the synthesis methods used and/or the groupings used in the synthesis, and how these affect the conclusions that can be drawn in relation to the original review question | 18-19 |  |

PRISMA=Preferred Reporting Items for Systematic Reviews and Meta-Analyses.

*If the information is not provided in the systematic review, give details of where this information is available (e.g., protocol, other published papers (provide citation details), or website (provide the URL)).

**Additional Table 2: Search Strategy**

**Ovid Embase**

1.	((abandon* or failed or shelved or "yet-to-be-pursued" or deprioritiz* or de-prioritiz* or re-discover* or rediscover* or repurpos* or re- purpos* or reposition* or re-position* or re-investigat* or reinvestigat*) adj3 (asset* or drug* or candidate* or pharma* or molecule* or compound* or biologic* or biopharmaceutical*)).tw,kw.

2.	exp drug repositioning/

3.	1 or 2

4.	(unapprov* or experiment* or investigat*).tw,kw.

5.	exp Drugs, Investigational/

6.	4 or 5

7.	3 and 6

**Ovid Medline**

1.	((abandon* or failed or shelved or "yet-to-be-pursued" or deprioritiz* or de-prioritiz* or re-discover* or rediscover* or repurpos* or re- purpos* or reposition* or re-position* or re-investigat* or reinvestigat*) adj3 (asset* or drug* or candidate* or pharma* or molecule* or compound* or biologic* or biopharmaceutical*)).tw,kw.

2.	exp drug repositioning/

3.	1 or 2

4.	(unapprov* or experiment* or investigat*).tw,kw.

5.	exp Drugs, Investigational/

6.	4 or 5

7.	3 and 6

**Web of Science Core Collection**

TS=((abandon* or failed or shelved or "yet-to-be-pursued" or deprioritiz* or "de-prioritiz*" or "re-discover*" or rediscover* or repurpos* or "re-purpos*" or reposition* or "re-position*" or "re-investigat*" or reinvestigat*) near/3 (asset* or drug* or candidate* or pharma* or molecule* or compound* or biologic* or biopharmaceutical*)) AND TS=(unapprov* or experiment* or investigat*)

**Scopus**

TITLE-ABS-KEY ( ( abandon*  OR  failed  OR  shelved  OR  "yet-to-be-pursued"  OR  deprioritiz*  OR  "de-prioritiz*"  OR  "re-discover*"  OR  rediscover*  OR  repurpos*  OR  "re-purpos*"  OR  reposition*  OR  "re-position*"  OR  "re-investigat*"  OR  reinvestigat* )  W/3  ( asset*  OR  drug*  OR  candidate*  OR  pharma*  OR  molecule*  OR  compound*  OR  biologic*  OR  biopharmaceutical* ) )  AND  TITLE-ABS-KEY ( unapprov*  OR  experiment*  OR  investigat* )

**Cochrane Library**

#1 ((abandon* or failed or shelved or "yet-to-be-pursued" or deprioritiz* or "de-prioritiz*" or "re-discover*" or rediscover* or repurpos* or "re-purpos*" or reposition* or "re-position*" or "re-investigat*" or reinvestigat*) near/3 (asset* or drug* or candidate* or pharma* or molecule* or compound* or biologic* or biopharmaceutical*)):ti,ab

#2 (unapprov* or experiment* or investigat*):ti,ab

#3 #1 and #2

**Pubmed**

((unapprov* [Title/Abstract] OR experiment* [Title/Abstract] OR investigat*)[Title/Abstract]) AND ((abandoned drug*[Title/Abstract] OR unapproved drug*[Title/Abstract] OR deprioritized drug*[Title/Abstract] OR failed drug*[Title/Abstract] OR repositioned drug*[Title/Abstract] OR reinvestigated drug*[Title/Abstract] or abandoned pharma*[Title/Abstract] OR unapproved pharma*[Title/Abstract] OR deprioritized pharma*[Title/Abstract] OR failed pharma*[Title/Abstract] OR repositioned pharma*[Title/Abstract] OR reinvestigated pharma*[Title/Abstract])

**Google Scholar**

1. “abandoned drug”
2. “deprioritized drug”
3. “shelved drug”

**Business Source Complete**
S1 TI ( unapprov* or experiment* or investigat* ) AND TI ( asset* or drug* or candidate* or pharma* or molecule* or compound* or biologic* or biopharmaceutical* ) AND TI ( abandon* or failed or shelved or "yet-to-be-pursued" or deprioritiz* or "de-prioritiz*" or "re-discover*" or rediscover* or repurpos* or "re-purpos*" or reposition* or "re-position*" or "re-investigat*" or reinvestigat* )

S2 AB ( abandon* or failed or shelved or "yet-to-be-pursued" or deprioritiz* or "de-prioritiz*" or "re-discover*" or rediscover* or repurpos* or "re-purpos*" or reposition* or "re-position*" or "re-investigat*" or reinvestigat* ) AND AB ( asset* or drug* or candidate* or pharma* or molecule* or compound* or biologic* or biopharmaceutical* ) AND AB ( unapprov* or experiment* or investigat*) )

S3 S1 OR S2

**Academic Source Complete**

S1 TI ( unapprov* or experiment* or investigat* ) AND TI ( asset* or drug* or candidate* or pharma* or molecule* or compound* or biologic* or biopharmaceutical* ) AND TI ( abandon* or failed or shelved or "yet-to-be-pursued" or deprioritiz* or "de-prioritiz*" or "re-discover*" or rediscover* or repurpos* or "re-purpos*" or reposition* or "re-position*" or "re-investigat*" or reinvestigat* )

S2 AB ( abandon* or failed or shelved or "yet-to-be-pursued" or deprioritiz* or "de-prioritiz*" or "re-discover*" or rediscover* or repurpos* or "re-purpos*" or reposition* or "re-position*" or "re-investigat*" or reinvestigat* ) AND AB ( asset* or drug* or candidate* or pharma* or molecule* or compound* or biologic* or biopharmaceutical* ) AND AB ( unapprov* or experiment* or investigat*) )

S3 S1 OR S2

**ABI/Informa**

noft(unapprov* or experiment* or investigat* ) AND noft(asset* or drug* or candidate* or pharma* or molecule* or compound* or biologic* or biopharmaceutical* ) AND noft(abandon* or failed or shelved or "yet-to-be-pursued" or deprioritiz* or "de-prioritiz*" or "re-discover*" or rediscover* or repurpos* or "re-purpos*" or reposition* or "re-position*" or "re-investigat*" or reinvestigat*)

**EconLit**

noft(unapprov* or experiment* or investigat* ) AND noft(asset* or drug* or candidate* or pharma* or molecule* or compound* or biologic* or biopharmaceutical* ) AND noft(abandon* or failed or shelved or "yet-to-be-pursued" or deprioritiz* or "de-prioritiz*" or "re-discover*" or rediscover* or repurpos* or "re-purpos*" or reposition* or "re-position*" or "re-investigat*" or reinvestigat*)

**Supplementary Table 3: Table of Excluded Studies with Reasons for Exclusion**

| **Year** | **First Author** | **Title** | **Journal** | **Exclusion Reason** |
| --- | --- | --- | --- | --- |
| 2018 | AbdelKhalek | Repurposing ebselen for decolonization of vancomycin-resistant enterococci (VRE) | PLoS ONE | No abandoned, shelved, or deprioritized drugs |
| 2012 | AbdulHameed | Exploring polypharmacology using a ROCS-based target fishing approach | Journal of Chemical Information and Modeling | No abandoned, shelved, or deprioritized drugs |
| 2019 | Acosta-Herrera | The potential role of genomic medicine in the therapeutic management of rheumatoid arthritis | Journal of Clinical Medicine | No abandoned, shelved, or deprioritized drugs |
| 2017 | Adams | The pharmacogenomics of severe traumatic brain injury | Pharmacogenomics | Approved drugs only |
| 2018 | Addeo | Learning from the past to design better trials in second-line treatment for mesothelioma patients | ecancermedicalscience | No abandoned, shelved, or deprioritized drugs |
| 2019 | Addeo | Association of industry and academic sponsorship with negative phase 3 oncology trials and reported outcomes on participant survival A Pooled Analysis | JAMA Network Open | No abandoned, shelved, or deprioritized drugs |
| 2017 | Agarwal | PI3-Kinase inhibitors represent a novel class of drug repurposing candidates to prevent/alleviate glucocorticoid-induced skin atrophy | Journal of Investigative Dermatology | Conference abstract |
| 2019 | Agrawal | The pursuit of excellence in new-drug development | McKinsey Insights | Conference abstract |
| 2016 | Ahmed | Repositioning of drugs using open-access data portal DTome: A test case with probenecid (Review) | International Journal of Molecular Medicine | Approved drugs only |
| 2019 | Aishah | Phenotypic approach to pharmacotherapy in the management of obstructive sleep apnoea | Current Opinion in Pulmonary Medicine | Approved drugs only |
| 2019 | Alberca | In silico Guided Drug Repurposing: Discovery of New Competitive and Non-competitive Inhibitors of Falcipain-2 | Frontiers of Chemistry | No abandoned, shelved, or deprioritized drugs |
| 2013 | Al-Ghananeem | Advances in brain targeting and drug delivery of anti-HIV therapeutic agents | Expert Opin Drug Deliv | No abandoned, shelved, or deprioritized drugs |
| 2019 | Alkhilaiwi | High-throughput screening identifies candidate drugs for the treatment of recurrent respiratory papillomatosis | Papillomavirus Research | No abandoned, shelved, or deprioritized drugs |
| 2017 | Almashat | Withholding Information on Unapproved Drug Marketing Applications: The Public Has a Right to Know | The Journal of Law, Medicine & Ethics | No abandoned, shelved, or deprioritized drugs |
| 2010 | Alving | Risks and rewards in creating partnerships among academia/industry/government to achieve public health outcomes | Clinical and Translational Science | Conference abstract |
| 2014 | Amelio | DRUGSURV: A resource for repositioning of approved and experimental drugs in oncology based on patient survival information | Cell Death and Disease | No abandoned, shelved, or deprioritized drugs |
| 2017 | Andresen | Drug Repurposing for the Treatment of Acute Myeloid Leukemia | Frontiers in Medicine | Approved drugs only |
| 2007 | Anonymous | Gene Logic Agrees to Sell Genomics Assets | Business Wire | No abandoned, shelved, or deprioritized drugs |
| 2008 | Anonymous | NEOPHARM REPORTS 2007 4TH QTR NET LOSS OF $1.1 MILLION | Biotech Financial Reports | No abandoned, shelved, or deprioritized drugs |
| 2012 | Anonymous | Revisiting 2007 And 2009 Disclosures | Chemical and Engineering News | No abandoned, shelved, or deprioritized drugs |
| 2016 | Antolin | Polypharmacology in precision oncology: Current applications and future prospects | Current Pharmaceutical Design | Approved drugs only |
| 2014 | Antoniu | Targeting 5-lipoxygenase-activating protein in asthma and chronic obstructive pulmonary disease | Expert Opinion on Therapeutic Targets | animal study |
| 2013 | Araki | Potential repurposing of oncology drugs for the treatment of Alzheimer's disease | BMC Medicine | Approved drugs only |
| 2019 | Armengol | A scientific task force to generate proof-of-concept data packages for clinical trials in pediatric cancers: The hepatoblastoma example | Cancer Research. Conference: American Association for Cancer Research Annual Meeting | Conference abstract |
| 2012 | Arrowsmith | Drug repositioning: the business case and current strategies to repurpose shelved candidates and marketed drugs | Drug Repositioning: Bringing New Life to Shelved Assets and Existing Drugs | Book chapter |
| 2013 | Asada | Analysis of new drugs whose clinical development and regulatory approval were hampered during their introduction in Japan | Journal of Clinical Pharmacy and Therapeutics | No abandoned, shelved, or deprioritized drugs |
| 2015 | Augustin | The wisdom of crowds and the repurposing of artesunate as an anticancer drug | ecancermedicalscience | Approved drugs only |
| 2017 | Austin | New therapeutic uses for existing drugs | Advances in Experimental Medicine and Biology | Book chapter |
| 2018 | Bahi | Drug-Target Interaction Prediction in Drug Repositioning Based on Deep Semi-Supervised Learning |  | Approved drugs only |
| 2020 | Ballas | The evolving pharmacotherapeutic landscape for the treatment of sickle cell disease | Mediterranean Journal of Hematology and Infectious Diseases | No abandoned, shelved, or deprioritized drugs |
| 2016 | Balmith | Ebola virus: A gap in drug design and discovery - experimental and computational perspective | Chemical Biology & Drug Design | No abandoned, shelved, or deprioritized drugs |
| 2016 | Barneh | Updates on drug-target network; facilitating polypharmacology and data integration by growth of DrugBank database | Briefings in bioinformatics | No abandoned, shelved, or deprioritized drugs |
| 2015 | Bellera | Knowledge-based drug repurposing: a rational approach towards the identification of novel medical applications of known drugs | Frontiers in computational chemistry | Book chapter |
| 2017 | Bellomo | High-content drug screening for rare diseases | Journal of Inherited Metabolic Disease | Approved drugs only |
| 2014 | Berg | Analysis of anakinra in primary human cell systems reveals an in vitro signature for skin-related side effects | Arthritis and Rheumatology | Conference abstract |
| 2012 | Bessoff | Improving therapeutics for the treatment of cryptosporidiosis using high throughput methods | American Journal of Tropical Medicine and Hygiene | Conference abstract |
| 2013 | Bharadwaj | Latrepirdine: molecular mechanisms underlying potential therapeutic roles in Alzheimer's and other neurodegenerative diseases | Transl Psychiatry | Approved drugs only |
| 2019 | BillettedeVillemeur | One lab, two firms, many possibilities: On R&D outsourcing in the biopharmaceutical industry | Journal of Health Economics | Duplicate |
| 2019 | Billones | Structure-Based Discovery of Inhibitors Against MurE in Methicillin-Resistant Staphylococcus aureus | Oriental Journal of Chemistry | No abandoned, shelved, or deprioritized drugs |
| 2016 | Bloom | Identification of iguratimod as an inhibitor of macrophage migration inhibitory factor (MIF) with steroid-sparing potential | Journal of Biological Chemistry | Approved drugs only |
| 2016 | Boileau | Identification of existing bioactive molecules that target acute myeloid leukemia stem cells | Experimental Hematology | Conference abstract |
| 2019 | Borba | Unveiling the Kinomes of Leishmania infantum and L. braziliensis Empowers the Discovery of New Kinase Targets and Antileishmanial Compounds | Computational and Structural Biotechnology Journal | No abandoned, shelved, or deprioritized drugs |
| 2008 | Boreham | Slivers of light in biotech sector gloom | Australasian Biotechnology | Insufficient data on repurposed, abandoned, or deprioritized drug |
| 2005 | Bower | From the `rhetoric of hopeÂ¿ to the `patient-active paradigm' - strategic positioning of pharmaceutical and biotechnology companies | Technology Analysis & Strategic Management | No abandoned, shelved, or deprioritized drugs |
| 2018 | Bridges | Cystic Fibrosis, Cystic Fibrosis Transmembrane Conductance Regulator and Drugs: Insights from Cellular Trafficking | Handbook of Experimental Pharmacology | Book chapter |
| 2014 | Broadstock | Latest treatment options for Alzheimer's disease, Parkinson's disease dementia and dementia with Lewy bodies | Expert Opinion on Pharmacotherapy | Approved drugs only |
| 2013 | Brundin | Developing new treatments founded on the basic science of Parkinson's disease | Journal of Parkinson's Disease | Conference abstract |
| 2015 | Bruning | Misfolded proteins: From little villains to little helpers in the fight against cancer | Frontiers in Oncology | No abandoned, shelved, or deprioritized drugs |
| 2003 | Butcher | Target discovery and validation in the post-genomic era | Neurochemical Research | No abandoned, shelved, or deprioritized drugs |
| 1990 | ByJerry | All in the Genes: New Medical Strategy Against Heart Disease Probes Inherited Flaws --- Scientists Begin to See Why Cholesterol Accumulates In Blood of Some People --- Mr. R's One Faulty Molecule | Wall Street Journal | No abandoned, shelved, or deprioritized drugs |
| 1998 | ByRhonda | Amgen Cancels Development Of Its MGDF | Wall Street Journal | No abandoned, shelved, or deprioritized drugs |
| 2014 | Cao | GWAS and drug targets | BMC Genomics | No abandoned, shelved, or deprioritized drugs |
| 2005 | Carley | Drug repurposing: Identify, develop and commercialize new uses for existing or abandoned drugs. Part I. 31 January - 1 February 2005, Philadelphia, PA, USA | IDrugs | Duplicate |
| 2005 | Carley | Drug repurposing: identify, develop and commercialize new uses for existing or abandoned drugs. Part II | Idrugs | Conference abstract |
| 2016 | Castangia | Old drugs, new therapies | Innovations in Pharmaceutical Technology | Approved drugs only |
| 2013 | Cavalla | Predictive methods in drug repurposing: Gold mine or just a bigger haystack? | Drug Discovery Today | Approved drugs only |
| 2017 | Ceskova | Novel treatment options in depression and psychosis | Clinical Therapeutics | No abandoned, shelved, or deprioritized drugs |
| 2013 | Chen | Drug repositioning by kernel-based integration of molecular structure, molecular activity, and phenotype data | PLoS ONE | No abandoned, shelved, or deprioritized drugs |
| 2014 | Chen | Pros and Cons of the Tuberculosis Drugome Approach â€“ An Empirical Analysis | PLoS ONE | Approved drugs only |
| 2014 | Chen | Pros and cons of the tuberculosis drugome approach - An empirical analysis | PLoS ONE | Approved drugs only |
| 2016 | Chen | Leveraging big data to transform target selection and drug discovery | Clinical Pharmacology and Therapeutics | No abandoned, shelved, or deprioritized drugs |
| 2017 | Cheng | Large-Scale Prediction of Drug-Target Interaction: a Data-Centric Review | AAPS Journal | Approved drugs only |
| 2012 | Chong | Repurposing drugs for tropical diseases: case studies and open-source screening initiatives |  | Book chapter |
| 2017 | Ciallella | In vivo phenotypic screening: clinical proof of concept for a drug repositioning approach | Drug Discovery Today: Technologies | Approved drugs only |
| 2016 | Clark | Preclinical Pain Research: Can We Do Better? | Anesthesiology | No abandoned, shelved, or deprioritized drugs |
| 2012 | Conn | A role for academic institutions in CNS drug discovery | International Journal of Neuropsychopharmacology | Conference abstract |
| 2017 | Corsello | Accelerating drug repurposing for cancer therapy using multiplexed viability assays | Cancer Research. Conference: American Association for Cancer Research Annual Meeting | Conference abstract |
| 2015 | DeCensi | Barriers to preventive therapy for breast and other major cancers and strategies to improve uptake | ecancermedicalscience | Approved drugs only |
| 2019 | deVillemeur | One lab, two firms, many possibilities: On R&D outsourcing in the biopharmaceutical industry | Journal of Health Economics | No abandoned, shelved, or deprioritized drugs |
| 2002 | DiMasi | The value of improving the productivity of the drug development process - Faster times and better decisions | PharmacoEconomics | No abandoned, shelved, or deprioritized drugs |
| 2003 | DiMasi | The price of innovation: New estimates of drug development costs | Journal of Health Economics | Insufficient data on repurposed, abandoned, or deprioritized drug |
| 2007 | DiMasi | Economics of new oncology drug development | Journal of Clinical Oncology | No abandoned, shelved, or deprioritized drugs |
| 2015 | Dodd | Future directions for pharmacotherapies for treatment-resistant bipolar disorder | Current Neuropharmacology | No abandoned, shelved, or deprioritized drugs |
| 2003 | Donnelly | Antimicrobial therapy to prevent or treat oral mucositis | Lancet Infectious Diseases | No abandoned, shelved, or deprioritized drugs |
| 2012 | Doudican | Rational design of a therapeutic program for multiple myeloma using a strategy of drug repurposing and combination therapy | Blood. Conference: 54th Annual Meeting of the American Society of Hematology, ASH | Conference abstract |
| 2010 | Dubus | Drug repurposing opportunities using in silico compound profiling | Drugs of the Future | Approved drugs only |
| 2019 | Elkhader | Drug repurposing engine fueled by diverse drug similarity data | Cancer Research. Conference: American Association for Cancer Research Annual Meeting | Conference abstract |
| 2014 | Eriksson | Repositioning of quinacrine for treatment of acute myeloid leukemia | Blood. Conference: 56th Annual Meeting of the American Society of Hematology, ASH | Conference abstract |
| 2019 | Farha | Drug repurposing for antimicrobial discovery | Nature Microbiology | Approved drugs only |
| 2016 | Federer | Big Data Mining and Adverse Event Pattern Analysis in Clinical Drug Trials | Assay and Drug Development Technologies | No abandoned, shelved, or deprioritized drugs |
| 1993 | Fenton | A unique drug activity search system | Journal of Information Science | No abandoned, shelved, or deprioritized drugs |
| 1985 | Finkelstein | Scientific Evidence and the Abandonment of Medical Technology: A Study of Eight Drugs | Research Policy | Approved drugs only |
| 2003 | Firn | Celltech's review gets a cool reception PHARMACEUTICALS: [LONDON 1ST EDITION] | Financial Times | No abandoned, shelved, or deprioritized drugs |
| 2012 | Frail | Opportunities and challenges associated with developing additional indications for clinical development candidates and marketed drugs | Drug Repositioning: Bringing New Life to Shelved Assets and Existing Drugs | Book chapter |
| 2013 | Frail | Drug repositioning through open innovation-fan industry perspective | Neuropsychopharmacology | Conference abstract |
| 2019 | Gao | PSPS: A pharmacological substances prediction system based on biomedical literature data |  | No abandoned, shelved, or deprioritized drugs |
| 2019 | Geisler | 2,4 Dinitrophenol as Medicine | Cells | No abandoned, shelved, or deprioritized drugs |
| 2018 | Giannuzzi | Failures to further developing orphan medicinal products after designation granted in Europe | Orphanet Journal of Rare Diseases. Conference: 9th European Conference on Rare Diseases and Orphan Products, ECRD | Conference abstract |
| 2019 | Glicksberg | Leveraging big data to transform drug discovery | Methods in Molecular Biology | Book chapter |
| 2018 | Goldstein | PregOMICS-Leveraging systems biology and bioinformatics for drug repurposing in maternal-child health | American Journal of Reproductive Immunology | Approved drugs only |
| 2018 | Goldstein | PregOMICSâ€”Leveraging systems biology and bioinformatics for drug repurposing in maternal-child health | American Journal of Reproductive Immunology | Approved drugs only |
| 2017 | Grabb | Challenges in developing drugs for pediatric CNS disorders: A focus on psychopharmacology | Progress in Neurobiology | No abandoned, shelved, or deprioritized drugs |
| 2015 | Graves | Tuberculosis vaccine development: Shifting focus amid increasing development challenges | Human Vaccines and Immunotherapeutics | No abandoned, shelved, or deprioritized drugs |
| 2013 | Grech | Targeted therapies in systemic lupus erythematosus | Lupus | No abandoned, shelved, or deprioritized drugs |
| 2002 | Green | Why do neuroprotective drugs that are so promising in animals fail in the clinic? An industry perspective | Clinical and Experimental Pharmacology and Physiology | No abandoned, shelved, or deprioritized drugs |
| 2019 | Griesenauer | CDEK: Clinical Drug Experience Knowledgebase | Database: The Journal of Biological Databases and Curation | Approved drugs only |
| 2017 | Grossman | Early Development Challenges for Drug Products Containing Nanomaterials | Aaps Journal | No abandoned, shelved, or deprioritized drugs |
| 2014 | Grover | Identification of novel therapeutics for complex diseases from genome-wide association data | BMC Medical Genomics | No abandoned, shelved, or deprioritized drugs |
| 2014 | Grunder | Challenges for novel treatment approaches in schizophrenia | International Journal of Neuropsychopharmacology | Conference abstract |
| 2017 | Guney | Investigating Side Effect Modules in the Interactome and Their Use in Drug Adverse Effect Discovery | Complex Networks Viii | Approved drugs only |
| 2019 | Hao | Open-source chemogenomic data-driven algorithms for predicting drug-target interactions | Briefings in bioinformatics | No abandoned, shelved, or deprioritized drugs |
| 2017 | Haslam | Learning disease relationships from clinical drug trials | Journal of the American Medical Informatics Association | No abandoned, shelved, or deprioritized drugs |
| 2013 | Hendrie | The failure of the antidepressant drug discovery process is systemic | Journal of Psychopharmacology | animal study |
| 2010 | Henkel | Challenges in the discovery and development of novel antibiotics - Will there be a new drug derived from natural sources? | Planta Medica. Conference: 7th Tannin Conference | Conference abstract |
| 2015 | Henry | Unleashing the power of comparative oncology models in nanomedicine research | European Journal of Nanomedicine | No abandoned, shelved, or deprioritized drugs |
| 2018 | Hintzsche | IMPACT web portal: Oncology database integrating molecular profiles with actionable therapeutics | BMC Medical Genomics | No abandoned, shelved, or deprioritized drugs |
| 2017 | Hoffer | Repositioning drugs for traumatic brain injury - N-acetyl cysteine and Phenserine | Journal of Biomedical Science | animal study |
| 2011 | Hong | Medication repurposing: New uses for old drugs | Journal of Pharmacy Technology | No abandoned, shelved, or deprioritized drugs |
| 2020 | Hong | Anti-inflammatory Strategies for Schizophrenia: A Review of Evidence for Therapeutic Applications and Drug Repurposing | Clinical Psychopharmacology & Neuroscience | Approved drugs only |
| 2014 | Huang | Drug repositioning discovery for early- and late-stage non-small-cell lung cancer | BioMed Research International | Approved drugs only |
| 2015 | Huang | A weighted and integrated drug-target interactome: drug repurposing for schizophrenia as a use case | BMC Systems Biology | Approved drugs only |
| 2020 | Huckle | Repositioning: Old drugs, new tricks | Regulatory Rapporteur | Approved drugs only |
| 2018 | Ianevski | Novel activities of safe-in-human broad-spectrum antiviral agents | Antiviral Research | No abandoned, shelved, or deprioritized drugs |
| 2019 | Ianevski | Expanding the activity spectrum of antiviral agents | Drug Discovery Today | No abandoned, shelved, or deprioritized drugs |
| 2019 | Idrus | In conversation with Cydan's Chris Adams and James McArthur | FierceBiotech | Insufficient data on repurposed, abandoned, or deprioritized drug |
| 2018 | Indelicato | Emerging therapeutics for the treatment of Friedreich's ataxia | Expert Opinion on Orphan Drugs | No abandoned, shelved, or deprioritized drugs |
| 2019 | Ion | Application of molecular framework-based data-mining method in the search for beta-secretase 1 inhibitors through drug repurposing | Journal of Biomolecular Structure and Dynamics | No abandoned, shelved, or deprioritized drugs |
| 2013 | Iorio | Transcriptional data: A new gateway to drug repositioning? | Drug Discovery Today | No abandoned, shelved, or deprioritized drugs |
| 2015 | Issa | RepurposeVS: A drug repurposing-focused computational method for accurate drug-target signature predictions | Combinatorial Chemistry and High Throughput Screening | Approved drugs only |
| 2020 | Issa | Machine and deep learning approaches for cancer drug repurposing | Seminars in Cancer Biology. | Approved drugs only |
| 2017 | Iwata | Elucidating the modes of action for bioactive compounds in a cell-specific manner by large-scale chemically-induced transcriptomics | Scientific Reports | No abandoned, shelved, or deprioritized drugs |
| 2007 | Jack | More open Sanofi EUROPEAN VIEW: [LONDON 1ST EDITION] | Financial Times | No abandoned, shelved, or deprioritized drugs |
| 2014 | Jahrling | Variola virus archives: a new century, a new approach | Lancet | No abandoned, shelved, or deprioritized drugs |
| 2018 | James | Ulimorelin, an Iv Ghrelin Agonist, Has Potent Prokinetic Effects in the Human Stomach but Not Colon: Why an Active Drug May Have Failed in Clinical Trials | Gastroenterology | Conference abstract |
| 2017 | Jardim | Factors associated with failure of oncology drugs in late-stage clinical development: A systematic review | Cancer Treatment Reviews | No abandoned, shelved, or deprioritized drugs |
| 2019 | Jiapaer | Identification of 2-fluoro palmitic acid as a potential therapeutic agent against glioblastoma | Neuro-Oncology | Conference abstract |
| 2012 | Jin | A novel method of transcriptional response analysis to facilitate drug repositioning for cancer therapy | Cancer Research | No abandoned, shelved, or deprioritized drugs |
| 2019 | Johnston | Repurposing drugs to treat L-DOPA-induced dyskinesia in Parkinson's disease | Neuropharmacology | Approved drugs only |
| 2020 | Kaneko | Drug Repositioning and Target Finding Based on Clinical Evidence | Biological & Pharmaceutical Bulletin | No abandoned, shelved, or deprioritized drugs |
| 2017 | Karatzas | Drug repurposing in idiopathic pulmonary fibrosis filtered by a bioinformatics-derived composite score | Scientific Reports | No abandoned, shelved, or deprioritized drugs |
| 2019 | Karatzas | An Application of Computational Drug Repurposing Based on Transcriptomic Signatures | Methods in Molecular Biology | Book chapter |
| 2005 | Kerber | Regulators boost efforts to track claims on drug progress | Knight Ridder Tribune Business News | No abandoned, shelved, or deprioritized drugs |
| 2017 | Khan | Emerging treatments for Alzheimer's disease for non-amyloid and non-tau targets | Expert Review of Neurotherapeutics | Approved drugs only |
| 2019 | Khosravi | Active repurposing of drug candidates for melanoma based on GWAS, PheWAS and a wide range of omics data | Molecular Medicine | Approved drugs only |
| 2011 | Kim | Butamben derivatives enhance BMP-2-stimulated commitment of C2C12 cells into osteoblasts with induction of voltage-gated potassium channel expression | Bioorganic and Medicinal Chemistry Letters | Approved drugs only |
| 2016 | Klug | Repurposing strategies for tropical disease drug discovery | Bioorganic and Medicinal Chemistry Letters | Approved drugs only |
| 2018 | Kozlina | Repurposing as a strategy for drug discovery. [Croatian] | Farmaceutski Glasnik | foreign language |
| 2016 | Kuang | Baseline Regularization for Computational Drug Repositioning with Longitudinal Observational Data | Ijcai | Approved drugs only |
| 2012 | Kubick | Buckthorn Wars and the Essence of Clinical Data | Applied Clinical Trials | No abandoned, shelved, or deprioritized drugs |
| 2019 | Kumar | Exploring the new horizons of drug repurposing: A vital tool for turning hard work into smart work | European Journal of Medicinal Chemistry | Approved drugs only |
| 2017 | Kuster | The target landscape of clinical kinase drugs | FEBS Journal | Conference abstract |
| 2018 | Kwok | Re-thinking Alzheimer's disease therapeutic targets using gene-based tests | Ebiomedicine | No abandoned, shelved, or deprioritized drugs |
| 2011 | Kwong | Discovery and development of telaprevir: an NS3-4A protease inhibitor for treating genotype 1 chronic hepatitis C virus | Nature Biotechnology | Approved drugs only |
| 2019 | Langreth | Confusing Alzheimer's Data Leaves Momentus Decision in FDA Hands | Bloomberg.com | Duplicate |
| 2018 | Lappin | Exploring compounds which have the potential to be repurposed into induction therapy for paediatric patients with acute myeloid leukaemia | Pediatric Blood and Cancer | Conference abstract |
| 2017 | Le | Drug respositioning by integrating known disease-gene and drug-target associations | Biomedical Research and Therapy | Conference abstract |
| 2018 | Le | Drug Repositioning by Integrating Known Disease-Gene and Drug-Target Associations in a Semi-supervised Learning Model | Acta Biotheoretica | Approved drugs only |
| 2016 | Lee | Repositioning Bevacizumab: A Promising Therapeutic Strategy for Cartilage Regeneration | Tissue Engineering - Part B: Reviews | Approved drugs only |
| 2015 | Lentini | Ebola therapy: Developing new drugs or repurposing old ones? | International Journal of Cardiology | Approved drugs only |
| 2011 | Li | A computational approach to finding novel targets for existing drugs | PLoS Computational Biology | No abandoned, shelved, or deprioritized drugs |
| 2012 | Li | Characterizing protein domain associations by Small-molecule ligand binding | Journal of Proteome Science & Computational Biology | Approved drugs only |
| 2016 | Li | Board Number: B1493 A multi-omic framework for evaluating the functional impact of the shared genetic landscape across common autoimmune diseases | Molecular Biology of the Cell | Conference abstract |
| 2016 | Li | A survey of current trends in computational drug repositioning | Briefings in bioinformatics | No abandoned, shelved, or deprioritized drugs |
| 2017 | Li | Identification of candidate drugs for the treatment of metastatic osteosarcoma through a subpathway analysis method | Oncology Letters | Approved drugs only |
| 2019 | Lightbody | A high-throughput drug screen reveals a novel compound class that significantly depletes IRF4 expression in multiple myeloma | Blood. Conference: 61st Annual Meeting of the American Society of Hematology, ASH | No abandoned, shelved, or deprioritized drugs |
| 2019 | Lin | A comprehensive evaluation of connectivity methods for L1000 data | Briefings in bioinformatics. | No abandoned, shelved, or deprioritized drugs |
| 2007 | Lindner | Clinical attrition due to biased preclinical assessments of potential efficacy | Pharmacology & Therapeutics | Insufficient data on repurposed, abandoned, or deprioritized drug |
| 2017 | Liu | Lessons Learned from Two Decades of Anticancer Drugs | Trends in Pharmacological Sciences | No abandoned, shelved, or deprioritized drugs |
| 2018 | Liu | Systematic polypharmacology and drug repurposing via an integrated L1000-based Connectivity Map database mining | Royal Society Open Science | Approved drugs only |
| 2019 | Liu | Three-Level Hepatotoxicity Prediction System Based on Adverse Hepatic Effects | Molecular Pharmaceutics | No abandoned, shelved, or deprioritized drugs |
| 2019 | Liu | Artificial intelligence and big data facilitated targeted drug discovery | Stroke and Vascular Neurology | Approved drugs only |
| 2015 | Lopes | The role of biomarkers in improving clinical trial success: A study of 1,079 oncology drugs | Journal of Clinical Oncology. Conference | Conference abstract |
| 2018 | Lopez-Ribot | Insights into new antibiofilm molecules | Medical Mycology | Conference abstract |
| 2017 | Louet | In silico model of the human ClC-Kb chloride channel: pore mapping, biostructural pathology and drug screening | Scientific Reports | Approved drugs only |
| 2004 | Loyd | Wyeth, Elan to report findings on experimental Alzheimer's vaccine | Knight Ridder Tribune Business News | No abandoned, shelved, or deprioritized drugs |
| 2018 | Lu | DR2DI: a powerful computational tool for predicting novel drug-disease associations | Journal of Computer-Aided Molecular Design | Approved drugs only |
| 2016 | Luo | Drug repositioning based on comprehensive similarity measures and Bi-Random walk algorithm | Bioinformatics (Oxford, England) | Approved drugs only |
| 2019 | Luo | Computational Drug Repositioning with Random Walk on a Heterogeneous Network | IEEE/ACM Transactions on Computational Biology and Bioinformatics | Approved drugs only |
| 2019 | Lyu | Shanghai Green Valley Wins China Approval for Alzheimer's Drug | Bloomberg.com | Insufficient data on repurposed, abandoned, or deprioritized drug |
| 2016 | Ma | Prediction of candidate drugs for treating pancreatic cancer by using a combined approach | PLoS ONE | Approved drugs only |
| 2019 | Ma | A comparative study of cluster detection algorithms in proteinâ€“protein interaction for drug target discovery and drug repurposing | Frontiers in Pharmacology | No abandoned, shelved, or deprioritized drugs |
| 2019 | Ma | A comparative study of cluster detection algorithms in protein-protein interaction for drug target discovery and drug repurposing | Frontiers in Pharmacology | No abandoned, shelved, or deprioritized drugs |
| 2008 | Mao | The toxicology of Clioquinol | Toxicology Letters | Approved drugs only |
| 2019 | Marais | Newer Drugs for Tuberculosis Prevention and Treatment in Children | Indian Journal of Pediatrics | No abandoned, shelved, or deprioritized drugs |
| 2017 | Martinez | Drugs in clinical development for the treatment of amyotrophic lateral sclerosis | Expert Opinion on Investigational Drugs | Approved drugs only |
| 2006 | Mayor | Cancer charity is to "borrow" candidate drugs shelved by companies | BMJ (Clinical research ed.) | Insufficient data on repurposed, abandoned, or deprioritized drug |
| 2017 | McCarthy | Drugs currently under investigation for the treatment of invasive candidiasis | Expert Opinion on Investigational Drugs | No abandoned, shelved, or deprioritized drugs |
| 2018 | McGarry | RESKO: Repositioning drugs by using side effects and knowledge from ontologies | Knowledge-Based Systems | No abandoned, shelved, or deprioritized drugs |
| 2019 | Mei | A multi-label learning framework for drug repurposing | Pharmaceutics | No abandoned, shelved, or deprioritized drugs |
| 2018 | Merrill | Lupus community panel proposals for optimising clinical trials: 2018 | Lupus Science & Medicine | No abandoned, shelved, or deprioritized drugs |
| 2011 | Michelson | Of lazarus and zombies: Looking for life after death in discontinued compounds | Neuropsychopharmacology | Conference abstract |
| 2019 | Miranda | Chagas disease treatment: From new therapeutic targets to drug discovery and repositioning | Current Medicinal Chemistry | No abandoned, shelved, or deprioritized drugs |
| 2017 | Mitchell | Profile: Centre for Clinical Brain Sciences, Edinburgh, UK | Lancet | Approved drugs only |
| 2013 | Mitka | Lithium May Decrease Suicide Risk | JAMA: Journal of the American Medical Association | No abandoned, shelved, or deprioritized drugs |
| 2014 | Mitsumoto | Clinical trials in amyotrophic lateral sclerosis: Why so many negative trials and how can trials be improved? | The Lancet Neurology | No abandoned, shelved, or deprioritized drugs |
| 2015 | Mokry | Mendelian randomisation applied to drug development in cardiovascular disease: A review | Journal of Medical Genetics | No abandoned, shelved, or deprioritized drugs |
| 2017 | Monacelli | Do Cancer Drugs Counteract Neurodegeneration? Repurposing for Alzheimer's Disease | Journal of Alzheimer's Disease | Approved drugs only |
| 2016 | Moosavinasab | 'RE:fine drugs': an interactive dashboard to access drug repurposing opportunities | Database : the journal of biological databases and curation | Approved drugs only |
| 2019 | Moridi | The assessment of efficient representation of drug features using deep learning for drug repositioning | BMC Bioinformatics | Approved drugs only |
| 2020 | Morofuji | Drug Development for Central Nervous System Diseases Using In Vitro Blood-brain Barrier Models and Drug Repositioning | Current pharmaceutical design. | Approved drugs only |
| 2016 | Mucke | Drug repurposing for vascular dementia: Overview and current developments | Future Neurology | Approved drugs only |
| 2014 | Mueller | Feasibility of genomics-enabled therapy for pediatric high-grade gliomas and diffuse pontine gliomas | Neuro-Oncology | Conference abstract |
| 2013 | Naveed | Structure-based protein-protein interaction networks and drug design | Quantitative Biology | Approved drugs only |
| 2013 | Nemeth | Calcimimetic and calcilytic drugs for treating bone and mineral-related disorders | Best Practice and Research: Clinical Endocrinology and Metabolism | Approved drugs only |
| 1988 | Newcombe | Drugs and AIDS: Radical proposals shelved | Mersey Drugs Journal | No abandoned, shelved, or deprioritized drugs |
| 2017 | Nishimura | Overcoming obstacles to drug repositioning in Japan | Frontiers in Pharmacology | No abandoned, shelved, or deprioritized drugs |
| 2019 | Nudelman | Biological Hallmarks of Cancer in Alzheimer's Disease | Molecular Neurobiology | Approved drugs only |
| 2015 | Nygaard | A phase Ib multiple ascending dose study of the safety, tolerability, and central nervous system availability of AZD0530 (saracatinib) in Alzheimer's disease | Alzheimer's Research and Therapy | Insufficient data on repurposed, abandoned, or deprioritized drug |
| 2019 | Okada | Entry Into New Therapeutic Areas: The Effect of Alliance on Clinical Trials | Therapeutic Innovation and Regulatory Science | No abandoned, shelved, or deprioritized drugs |
| 2015 | Omer | Explicit Drug Re-positioning: Predicting Novel Drugâ€“Target Interactions of the Shelved Molecules with QM/MM Based Approaches | Advances in Protein Chemistry and Structural Biology | Book chapter |
| 2015 | Oprea | Computational and practical aspects of drug repositioning | Assay and Drug Development Technologies | Approved drugs only |
| 2018 | Pantziarka | Omics-driven drug repurposing as a source of innovative therapies in rare cancers | Expert Opinion on Orphan Drugs | Approved drugs only |
| 2015 | Patsopoulos | Translational genetics to identify the druggable space of neurological diseases | Neurology. Conference: 67th American Academy of Neurology Annual Meeting, AAN | Conference abstract |
| 2014 | Patterson | Nitro drugs for the treatment of trypanosomatid diseases: Past, present, and future prospects | Trends in Parasitology | No abandoned, shelved, or deprioritized drugs |
| 2013 | Penel | Activity endpoints reported in soft tissue sarcoma phase II trials: Quality of reported endpoints and correlation with overall survival | Critical Reviews in Oncology/Hematology | No abandoned, shelved, or deprioritized drugs |
| 2011 | Pennings | Evaluating pharmaceutical R&D under technical and economic uncertainty | European Journal of Operational Research | No abandoned, shelved, or deprioritized drugs |
| 2014 | Perkins | Improving efficiency of initial tests for efficacy in smoking cessation drug discovery | Expert Opinion on Drug Discovery | Approved drugs only |
| 2003 | Peuckmann | [Novel potential uses of thalidomide in the management of pain? A review of the literature] | Schmerz | Approved drugs only |
| 2016 | Piddock | Ask the experts: How to curb antibiotic resistance and plug the antibiotics gap? | Future Medicinal Chemistry | Approved drugs only |
| 2013 | Pinault | Zafirlukast inhibits complexation of Lsr2 with DNA and growth of Mycobacterium tuberculosis | Antimicrobial Agents & Chemotherapy | No abandoned, shelved, or deprioritized drugs |
| 2016 | Polonen | Molecular classification and comparative analysis of acute myeloid leukemia multi-omics data using the interactive hemap resource | Haematologica | Conference abstract |
| 2015 | Purow | Molecular pathways: Targeting diacylglycerol kinase alpha in cancer | Clinical Cancer Research | No abandoned, shelved, or deprioritized drugs |
| 2012 | Qu | Applications of Connectivity Map in drug discovery and development | Drug Discovery Today | Approved drugs only |
| 2014 | Qu | Integrative clinical transcriptomics analyses for new therapeutic intervention strategies: A psoriasis case study | Drug Discovery Today | No abandoned, shelved, or deprioritized drugs |
| 2018 | Rahman | Children's brain tumour drug delivery consortium | Neuro-Oncology | Conference abstract |
| 2018 | Rai | Systems biology: A powerful tool for drug development | Current Topics in Medicinal Chemistry | No abandoned, shelved, or deprioritized drugs |
| 2015 | Rastegar-Mojarad | Toward a Complete Database of Drug Repurposing Candidates Extracted from Social Media, Biomedical Literature, and Genetic Data |  | Conference abstract |
| 2015 | Ritchie | Cancer research UK centre for drug development: Translating 21st-century science into the cancer medicines of tomorrow | Drug Discovery Today | No abandoned, shelved, or deprioritized drugs |
| 2011 | Robertson | Drug discovery for neglected tropical diseases at the Sandler Center | Future Medicinal Chemistry | Approved drugs only |
| 2017 | Rogaeva | Drug re-positioning for alzheimer's disease based on systematic 'omics' data minding | Neurodegenerative Diseases | Conference abstract |
| 2019 | Rommer | Repurposing multiple sclerosis drugs: a review of studies in neurological and psychiatric conditions | Drug Discovery Today | Approved drugs only |
| 1996 | Rowe | Camptothecins: New enthusiasm for an old drug | Lancet | No abandoned, shelved, or deprioritized drugs |
| 2011 | Rowe | Two neglected diseases share similar parasites and suffer the same diagnostic and treatment problems | Chemical and Engineering News | No abandoned, shelved, or deprioritized drugs |
| 2016 | Ru | A proof-of-concept study of mining social media to reposition drugs | Value in Health | Conference abstract |
| 2020 | Rubin | The frontiers of addressing antibiotic resistance in Neisseria gonorrhoeae | Translational Research. | Approved drugs only |
| 1998 | Rundle | Amgen abandons development work on clotting drug | Wall Street Journal - Eastern Edition | No abandoned, shelved, or deprioritized drugs |
| 2019 | Sadeghi | An Analytical Review of Computational Drug Repurposing | IEEE/ACM transactions on computational biology and bioinformatics. | Approved drugs only |
| 2019 | Sadeghi | RCDR: A Recommender Based Method for Computational Drug Repurposing | 2019 Ieee 5th Conference on Knowledge Based Engineering and Innovation | Approved drugs only |
| 2016 | Sahu | Computational Drug Repositioning: A Lateral Approach to Traditional Drug Discovery? | Current Topics in Medicinal Chemistry | No abandoned, shelved, or deprioritized drugs |
| 2018 | Saiz | Host-Directed Antivirals: A Realistic Alternative to Fight Zika Virus | Viruses | Approved drugs only |
| 2019 | Salas-Sarduy | Target-based screening of the chagas box: Setting up enzymatic assays to discover specific inhibitors across bioactive compounds | Current Medicinal Chemistry | No abandoned, shelved, or deprioritized drugs |
| 2016 | Salomao | Stairway to heaven or hell? Perspectives and limitations of chagas disease chemotherapy | Current Topics in Medicinal Chemistry | No abandoned, shelved, or deprioritized drugs |
| 2014 | Sanchez-Jimenez | Biocomputational resources useful for drug discovery against compartmentalized targets | Current Pharmaceutical Design | No abandoned, shelved, or deprioritized drugs |
| 2014 | SanLucas | Drug repositioning with a bioinformatics platform that integrates the TCGA, cMap and CCLE | Cancer Research. | Conference abstract |
| 2014 | Sanseau | Genetics and drug repositioning | Drug Metabolism Reviews | Conference abstract |
| 2014 | Savva | Opt-Out Options in New Product Co-development Partnerships | Production and Operations Management | No abandoned, shelved, or deprioritized drugs |
| 2019 | Saye | Amino acid and polyamine membrane transporters in Trypanosoma cruzi: Biological function and evaluation as drug targets | Current Medicinal Chemistry | Approved drugs only |
| 2020 | Scherman | Drug repositioning for rare diseases: Knowledge-based success stories | Therapie. | Approved drugs only |
| 2016 | Schmidt | Cystic fibrosis transmembrane conductance regulator modulators in cystic fibrosis: Current perspectives | Clinical Pharmacology: Advances and Applications | Approved drugs only |
| 2005 | Schuster | Why drugs fail - A study on side effects in new chemical entities | Current Pharmaceutical Design | Approved drugs only |
| 2002 | Sekine | Relationship between objective responses in phase I trials and potential efficacy of non-specific cytotoxic investigational new drugs | Annals of Oncology | No abandoned, shelved, or deprioritized drugs |
| 2006 | Shah | Can pharmacogenetics help rescue drugs withdrawn from the market? |  | Approved drugs only |
| 2015 | Shamas-Din | Drug discovery in academia | Experimental Hematology | No abandoned, shelved, or deprioritized drugs |
| 2018 | Shameer | Systematic analyses of drugs and disease indications in RepurposeDB reveal pharmacological, biological and epidemiological factors influencing drug repositioning | Briefings in bioinformatics | Approved drugs only |
| 2017 | Sharp | Development of repurposed drugs for neuroscience: An example from academia | International Journal of Nutrition, Pharmacology, Neurological Diseases | Conference abstract |
| 2011 | Shen | Designs can increase efficiency in psychiatric drug development: A case study | Innovations in Clinical Neuroscience | No abandoned, shelved, or deprioritized drugs |
| 2018 | Shen | Quantitative high-throughput phenotypic screening of pediatric cancer cell lines identifies multiple opportunities for drug repurposing | Oncotarget | No abandoned, shelved, or deprioritized drugs |
| 2018 | Singh | Triple monoamine reuptake inhibitor of dopamine, norepinephrine and serotonin treatment of obesity | Drugs of the Future | No abandoned, shelved, or deprioritized drugs |
| 2020 | Singh | Recent trends in management of Alzheimer's disease: Current therapeutic options and drug repurposing approaches | Current neuropharmacology. | No abandoned, shelved, or deprioritized drugs |
| 2007 | Slatko | Court upholds policy on experimental drugs | Med Ad News | Insufficient data on repurposed, abandoned, or deprioritized drug |
| 2019 | Southall | The use or generation of biomedical data and existing medicines to discover and establish new treatments for patients with rare diseases-recommendations of the IRDiRC Data Mining and Repurposing Task Force | Orphanet Journal of Rare Diseases | No abandoned, shelved, or deprioritized drugs |
| 2016 | Spellberg | Regulatory Pathways for New Antimicrobial Agents: Trade-offs to Keep the Perfect From Being the Enemy of the Good | Clinical Pharmacology and Therapeutics | No abandoned, shelved, or deprioritized drugs |
| 2019 | Spiekerkoetter | New and emerging therapies for pulmonary arterial hypertension | Annual Review of Medicine | No abandoned, shelved, or deprioritized drugs |
| 2006 | Splawinski | Evaluation of drug toxicity in clinical trials | Science and Engineering Ethics | No abandoned, shelved, or deprioritized drugs |
| 2016 | Stern | A Perspective on Implementing a Quantitative Systems Pharmacology Platform for Drug Discovery and the Advancement of Personalized Medicine | Journal of Biomolecular Screening | No abandoned, shelved, or deprioritized drugs |
| 2004 | Strovel | Early Drug Discovery and Development Guidelines: For Academic Researchers, Collaborators, and Start-up Companies | Eli Lilly & Company and the National Center for Advancing Translational Sciences | Approved drugs only |
| 2018 | Subudhi | Current strategies for inhibition of Chikungunya infection | Viruses | No abandoned, shelved, or deprioritized drugs |
| 2019 | Sun | Overcoming Multidrug-Resistance in Bacteria with a Two-Step Process to Repurpose and Recombine Established Drugs | Analytical Chemistry | No abandoned, shelved, or deprioritized drugs |
| 2014 | Talekar | Tumor aerobic glycolysis: new insights into therapeutic strategies with targeted delivery | Expert Opinion on Biological Therapy | No abandoned, shelved, or deprioritized drugs |
| 2016 | Talevi | The importance of drug repurposing in the field of antiepileptic drug development | Antiepileptic Drug Discovery | Book chapter |
| 2012 | Tamargo | Cardiovascular risk and drugs | Basic and Clinical Pharmacology and Toxicology | Conference abstract |
| 2018 | Tanoli | Interactive visual analysis of drug-target interaction networks using Drug Target Profiler, with applications to precision medicine and drug repurposing | Briefings in Bioinformatics | No abandoned, shelved, or deprioritized drugs |
| 2017 | Tian | Repurposing Lesogaberan to Promote Human Islet Cell Survival and Î² -Cell Replication | Journal of Diabetes Research | animal study |
| 1987 | Tosch | DO PATIENTS BENEFIT FROM A FAILED EXPERIMENTAL ANTIEPILEPTIC DRUG STUDY | Epilepsia | Conference abstract |
| 2014 | Toumi | Drug repurposing as an efficient strategy in drug development-example of CNS area | Value in Health | Conference abstract |
| 2017 | Tseng | Accelerating prediction of pediatric and rare cancer vulnerabilities using next-generation cancer models | Cancer Research. Conference: American Association for Cancer Research Annual Meeting | Conference abstract |
| 2017 | Tsuji | AI technology based drug discovery method with protein interaction network for cancer drug repositioning | Cancer Science | Conference abstract |
| 2017 | Unger | Current Care and Investigational Therapies in Achondroplasia | Current Osteoporosis Reports | Approved drugs only |
| 2013 | Vaduganathan | The disconnect between phase II and phase III trials of drugs for heart failure | Nature Reviews Cardiology | No abandoned, shelved, or deprioritized drugs |
| 2020 | Vanhaelen | Web-based tools for drug repurposing: successful examples of collaborative research | Current medicinal chemistry. | Approved drugs only |
| 2015 | Veljkovic | Virtual screen for repurposing approved and experimental drugs for candidate inhibitors of EBOLA virus infection | F1000Research | No abandoned, shelved, or deprioritized drugs |
| 2019 | Vijayakumar | LeishInDB: A web-accessible resource for small molecule inhibitors against Leishmania sp | Acta Tropica | No abandoned, shelved, or deprioritized drugs |
| 2008 | Vincenti | What's next in the pipeline | American Journal of Transplantation | No abandoned, shelved, or deprioritized drugs |
| 2016 | Vitali | A network-based data integration approach to support drug repurposing and multi-Target therapies in triple negative breast cancer | PLoS ONE | No abandoned, shelved, or deprioritized drugs |
| 2019 | Vitiello | Teaching an old molecule new tricks: Drug repositioning for duchenne muscular dystrophy | International Journal of Molecular Sciences | Approved drugs only |
| 2008 | Wang | Myriad Drug Fails to Fight Alzheimer's | Wall Street Journal - Eastern Edition | No abandoned, shelved, or deprioritized drugs |
| 2013 | Wang | Clinical and regulatory features of drugs not initially approved by the FDA | Clinical Pharmacology and Therapeutics | No abandoned, shelved, or deprioritized drugs |
| 2019 | Wang | Predicting associations among drugs, targets and diseases by tensor decomposition for drug repositioning | BMC Bioinformatics | Approved drugs only |
| 2019 | Wang | A Gated Recurrent Unit Model for Drug Repositioning by Combining Comprehensive Similarity Measures and Gaussian Interaction Profile Kernel |  | Approved drugs only |
| 2009 | Weintraub | Tough Decisions on Osteoporosis Drugs | Business Week (Online) | No abandoned, shelved, or deprioritized drugs |
| 2019 | Weissenrieder | Cancer and the Dopamine D2 Receptor: A Pharmacological Perspective | Journal of Pharmacology and Experimental Therapeutics | No abandoned, shelved, or deprioritized drugs |
| 2000 | Wiendl | [Multiple sclerosis. Current review of failed and discontinued clinical trials of drug treatment] | Nervenarzt | Insufficient data on repurposed, abandoned, or deprioritized drug |
| 2014 | Wilkins | Pulmonary hypertension: the value of experimental medicine in new drug development | Pulmonary Circulation | Approved drugs only |
| 2015 | Wilkinson | In vitro screening for drug repositioning | Journal of Biomolecular Screening | No abandoned, shelved, or deprioritized drugs |
| 2016 | Winblad | What will be the best treatment for alzheimer disease? | Neurobiology of Aging | Conference abstract |
| 2018 | Wood | Identifying and accelerating potential new drug therapies for pediatric atypical teratoid rhabdoid tumors (ATRTS) through drug repurposing | Neuro-Oncology | Conference abstract |
| 2013 | Wu | Computational drug repositioning through heterogeneous network clustering | BMC Systems Biology | Approved drugs only |
| 2016 | Wu | In silico prediction of chemical mechanism of action via an improved network-based inference method | British Journal of Pharmacology | No abandoned, shelved, or deprioritized drugs |
| 2017 | Wu | DrugSig: A resource for computational drug repositioning utilizing gene expression signatures | PLoS ONE | No abandoned, shelved, or deprioritized drugs |
| 2018 | Xiong | Refocusing neuroprotection in cerebral reperfusion era: New challenges and strategies | Frontiers in Neurology | No abandoned, shelved, or deprioritized drugs |
| 2019 | Xuan | Drug repositioning through integration of prior knowledge and projections of drugs and diseases | Bioinformatics (Oxford, England) | Approved drugs only |
| 2018 | Xue | Review of Drug Repositioning Approaches and Resources | International Journal of Biological Sciences | Approved drugs only |
| 2019 | Yadavalli | Repurposed Drugs in Treating Glioblastoma Multiforme: Clinical Trials Update | Cancer Journal (United States) | Approved drugs only |
| 2017 | Yan | SDTRLS: Predicting Drug-Target Interactions for Complex Diseases Based on Chemical Substructures | Complexity | No abandoned, shelved, or deprioritized drugs |
| 2019 | Yang | Mining heterogeneous network for drug repositioning using phenotypic information extracted from social media and pharmaceutical databases | Artificial Intelligence in Medicine | Approved drugs only |
| 2005 | Yasui | Thalidomide as an immunotherapeutic agent: the effects on neutrophil-mediated inflammation | Current Pharmaceutical Design | Approved drugs only |
| 2012 | Yogesh | Bioavailability: A pharmaceutical importance in new drug development | Research Journal of Pharmaceutical, Biological and Chemical Sciences | No abandoned, shelved, or deprioritized drugs |
| 2020 | Zahora¡nszky-Kohalmi | SmartGraph: A network pharmacology investigation platform | Journal of Cheminformatics | No abandoned, shelved, or deprioritized drugs |
| 2013 | Zhang | Computational drug repositioning by ranking and integrating multiple data sources |  | Book chapter |
| 2014 | Zhang | Towards drug repositioning: a unified computational framework for integrating multiple aspects of drug similarity and disease similarity | Amia .. | No abandoned, shelved, or deprioritized drugs |
| 2016 | Zhang | Drug repositioning for Alzheimer's disease based on systematic 'omics' data mining | PLoS ONE | No abandoned, shelved, or deprioritized drugs |
| 2017 | Zhang | Computational drug repositioning using collaborative filtering via multi-source fusion | Expert Systems with Applications | Approved drugs only |
| 2019 | Zhang | Sample Size Considerations for a Phase III Clinical Trial with Diluted Treatment Effect | Statistics in Biopharmaceutical Research. | No abandoned, shelved, or deprioritized drugs |
| 2013 | Zhao | Novel modeling of cancer cell signaling pathways enables systematic drug repositioning for distinct breast cancer metastases | Cancer Research | No abandoned, shelved, or deprioritized drugs |
| 2016 | Zhao | Drug repurposing to target Ebola virus replication and virulence using structural systems pharmacology | BMC Bioinformatics | No abandoned, shelved, or deprioritized drugs |
| 2019 | Zhao | EK-DRD: A Comprehensive Database for Drug Repositioning Inspired by Experimental Knowledge | Journal of Chemical Information and Modeling | Approved drugs only |
| 2018 |  | Atriva Therapeutics GmbH - Pharmaceuticals &amp; Healthcare - Deals and Alliances Profile |  | No abandoned, shelved, or deprioritized drugs |
| 2015 |  | House Committee To Mark Up 21st Century Cures Package |  | Insufficient data on repurposed, abandoned, or deprioritized drug |
| 2005 |  | No Flu Shot Shortage Health and ... [Derived headline] | Investor's Business Daily | No abandoned, shelved, or deprioritized drugs |
| 2014 |  | Genmab inks research deal for DuoBody, HexaBody antibody technologies | M2 Pharma | Insufficient data on repurposed, abandoned, or deprioritized drug |
| 2003 |  | Development plans for new drug abandoned | AIDS Patient Care and STDs | Insufficient data on repurposed, abandoned, or deprioritized drug |
| 2007 |  | Client of the Month Club | CQ Weekly | Insufficient data on repurposed, abandoned, or deprioritized drug |
| 2019 |  | Atriva Therapeutics GmbH - Pharmaceuticals &amp; Healthcare - Deals and Alliances Profile |  | No abandoned, shelved, or deprioritized drugs |
| 2012 |  | TARGACEPT INC | MondayMorning | No abandoned, shelved, or deprioritized drugs |
| 2007 |  | Merck: Is Mercks Medicine Working? | Business Week (Online) | No abandoned, shelved, or deprioritized drugs |
| 1974 |  | FDA ABOUT-FACE SETS OFF NEW ROUND OF DMSO RESEARCH | Medical World News | Approved drugs only |
| 2005 |  | KineMed Announces Award of SBIR Phase I Grant for Program in Neurobiology | PR Newswire | No abandoned, shelved, or deprioritized drugs |
| 2014 |  | Virginia Biosciences Health Research to Fund 4 Private-Public Research Collaborations | Health & Beauty Close - Up | Approved drugs only |
| 2007 |  | Pharmaceutical Company Round-up | PharmaWatch: Monthly Review | No abandoned, shelved, or deprioritized drugs |
| 2012 |  | Research and Markets: Oncology Drug Pathway Analyzer (Premium Version) | Business Wire | No abandoned, shelved, or deprioritized drugs |
| 2004 |  | Helix BioPharma Corp.: DOS47 Update | Business Wire | No abandoned, shelved, or deprioritized drugs |
| 2019 |  | Resurrection: Deploying drugs for new purposes holds great promise | Economist | No abandoned, shelved, or deprioritized drugs |

**Additional Table 4: Table of Included Studies**

| **Year** | **First Author** | **Title** | **Terms used** | **Reasons Drugs were Shelved** | **Barriers** | **Facilitators** | **Medical Diseases** | **Publication Type** | **Oxford**  **Scale Evidence** |
| --- | --- | --- | --- | --- | --- | --- | --- | --- | --- |
| 2013 | Allarakhia | Open-source approaches for the repurposing of existing or failed candidate drugs: Learning from and applying the lessons across diseases | Repurposing |  | Patent and IP issues, Low ROI potential | Collaborative initiatives, Compound libraries and/or drug-disease databases | Neglected tropical diseases and orphan diseases | Review | - |
| 2007 | Andrew | AstraZeneca likely to abandon drug |  | Efficacy |  |  |  | Press release/  news article | 5 |
| 2015 | Anonymous | New indications and a sense of (re)purpose | Repositioning, Repurposing, Resurrecting |  | Bias | Collaborative initiatives | Alzheimer's disease | Expert Opinion/  editorial | 5 |
| 2017 | Anonymous | Lilly, rivals not backing away from pricey Alzheimer's studies | Abandoned, Failed | Efficacy | Low ROI potential |  | Alzheimer's disease | Press release/  news article | 5 |
| 2014 | Auer | Translational research and efficacy of biologics in Crohn's disease: a cautionary tale | Failed | Safety issues, Efficacy |  |  | Crohn's Disease | Review | - |
| 2013 | Becker | Fire in the ashes: Can failed Alzheimer's disease drugs succeed with second chances? | Redevelopment | Efficacy, Practices in development | Flawed Methodology, Bias |  | Neurodegenerative disease, Alzheimer's disease | Article | 3 |
| 2012 | Becker | Was phenserine a failure or were investigators mislead by methods? |  | Efficacy, Trials Failing Drugs |  |  | Alzheimer's disease | Article | 3 |
| 2010 | Becker | Lost in Translation: Neuropsychiatric Drug Development | Failed | Strategic reasons, Development and Design |  |  | Neuropsychiatric disease | Review | - |
| 2020 | Bellera | In silico Drug Repositioning for Chagas Disease | Repositioning, Repurposing, Rescue |  | Access to data | Compound libraries and/or drug-disease databases, Use of computational methods/systematic screening methods | Neglected tropical disease | Review | - |
| 2005 | Bishop | Repurposing companies experiment with unintended uses for pharmaceuticals | Repositioning, Repurposing, Reprofiling, Rescue | Efficacy,  Strategic reasons |  | Repositioning companies |  | Press release/  news article | 5 |
| 2012 | Bisson | Drug repurposing in chemical genomics: Can we learn from the past to improve the future? | Repurposing |  | Patent and IP issues, Low ROI potential, Access to data, Risk | Collaborative initiatives, Compound libraries and/or drug-disease databases, Use of computational methods/systematic screening methods | Rare and neglected diseases | Article | 4 |
| 2014 | Buonansegna | Pharmaceutical new product development: Why do clinical trials fail? | Abandoned | Safety issues,  Efficacy,  Strategic reasons,  Lack of Experience, Flaws in Methods |  |  |  | Article | 4 |
| 2018 | Cha | Drug repurposing from the perspective of pharmaceutical companies | Repurposing | Safety issues,  Efficacy | Patent and IP issues, Low ROI potential | Collaborative initiatives, Compound libraries and/or drug-disease databases, Use of computational methods/systematic screening methods, Regulatory modifications |  | Review | - |
| 2013 | Chesbrough | Recovering abandoned compounds through expanded external IP licensing |  | Safety issues, Efficacy, Strategic reasons, Evaluation errors, Industry consolidation | Bias, Patent and IP issues, Low ROI potential, Liability |  |  | Article | 4 |
| 2015 | Coaker | Neglected tropical disease research: rethinking the drug discovery model | Repositioning, Repurposing, Reprofiling, Rescue, Redirecting, Therapeutic Switching |  | Bias, Out of scope/ lack of expertise, Patent and IP issues, Low ROI potential | Collaborative initiatives, Compound libraries and/or drug-disease databases, Regulatory modifications | Neglected Tropical Diseases | Expert Opinion/  Editorial | 5 |
| 2011 | Collins | Mining for therapeutic gold | Repurposing, Rescue | Efficacy | Patent and IP issues, Liability | Collaborative initiatives, Use of computational methods/systematic screening methods, Master access, Central access point, Data and resource availability | Rare disease | Comment | 5 |
| 2019 | Cortez | Biogen resumes Alzheimer's studies to offer controversial drug |  | Efficacy |  |  | Alzheimer's disease | Press release/  News article | 5 |
| 2017 | Crow | J&J boss’s long quest ends with $30bn Actelion deal |  | Strategic reasons | Low ROI potential, High cost | Acquisition deal structure |  | Press release/ New Article | 5 |
| 2018 | Challener | Can Artificial Intelligence Take the Next Step for Drug Repositioning | Repositioning |  |  | Use of computational methods/systematic screening methods |  | Expert Opinion/ Editorial | 5 |
| 2015 | Daniel | Improving pharmaceutical innovation by building a more comprehensive database on drug development and use | Abandoned |  |  | Collaborative initiatives, Compound libraries and/or drug-disease databases |  | Expert Opinion/ Editorial | 5 |
| 2017 | de Villiers | The quest for new drugs to prevent osteoporosis-related fractures |  | Safety issues, Efficacy |  |  |  | Review | - |
| 2011 | Von Eichborn | PROMISCUOUS: A database for network-based drug-repositioning | Repositioning | Safety issues, Efficacy | Access to data | Compound libraries and/or drug-disease databases |  | Article |  |
| 2010 | Empfield | Lessons learned from candidate drug attrition |  | Safety issues, Efficacy |  |  |  | Review | - |
| 2006 | Feemster | Rescue Squad | Repositioning, Shelved, Discontinued, Failed, Dead drugs | Safety issues, Efficacy, Strategic reasons | Patent and IP issues, Low ROI potential | Collaborative initiatives, Use of computational methods/systematic screening methods, Strong relationships, Pipeline crisis in pharma |  | Press release/ New Article | 5 |
| 2020 | Feijoo | Key indicators of phase transition for clinical trials through machine learning | Failed | Trial Design |  |  |  | Review | - |
| 2020 | Fetro | Drug repurposing in rare diseases: Myths and reality | Repositioning, Repurposing, Reprofiling, Rescue, Rediscovery, Retasking |  | Patent and IP issues, Risk, Time consuming, Expensive | Incentives | Rare diseases | Article | 5 |
| 2004 | Firn | Roche drops trials of new HIV drug |  | Strategic reasons |  |  |  | Press release/ New Article | 5 |
| 2015 | Frail | Pioneering government-sponsored drug repositioning collaborations: Progress and learning | Repositioning, Repurposing, Reprofiling, Rescue, Indications discovery |  | Bias, Out of scope/ lack of expertise, Patent and IP issues, Low ROI potential, Liability, Cost, Personnel | Collaborative initiatives |  | Expert Opinion/ Editorial | 5 |
| 2015 | Friedhoff | Lost interest for existing compounds: New boosts | Re-initiating development | Safety issues | Patent and IP issues | Collaborative initiatives, Compound libraries and/or drug-disease databases, Data sharing consortia |  | Article | 5 |
| 2017 | Giannuzzi | Failures to further developing orphan medicinal products after designation granted in Europe | Abandoned | Efficacy, Strategic reasons, Drug Competition, Inactive Companies |  | Tax incentives, Fee waivers, Market exclusivity, Public funding | Rare diseases | Article | 3 |
| 2005 | Griel | Pfizer Drops 2 Potential Drugs |  | Efficacy, Strategic reasons | Bias, Cost |  |  | Press release/ New Article | 5 |
| 2019 | Griffin | Biogen Surges as Momentum for Alzheimer's Treatment Revives Hope | Reviving, Abandoned | Efficacy |  | Re-analysis of data | Alzheimer's disease | Press release/ New Article | 5 |
| 2014 | Grundy | Reconfiguring drug discovery through innovative partnerships |  | Efficacy, Chronic and Multifactorial conditions |  | Collaborative initiatives, Open-source drug discovery | Alzheimer's disease | Press release/ New Article | 5 |
| 2015 | Gunther | Big pharma opens up abandoned drugs | Repurposing, Abandoned, Deprioritized, Stalled |  |  | Collaborative initiatives |  | Newsletter | 5 |
| 2018 | Guo | Is it time for a paradigm shift in drug research and development in endometriosis/adenomyosis? | Repurposing, Abandoned | Safety issues, Efficacy, Strategic reasons |  |  | Endometriosis and adenomyosis | Article | - |
| 2019 | Hayes | Compound asset sharing initiatives between pharmaceutical companies, funding bodies, and academia: Learnings and Successes British Pharmacological Society Journals | Repurposing, |  | Patent and IP issues, Legal Agreements | Collaborative initiatives | CNS | Commentary | 5 |
| 2015 | Hayes | The European College of Neuropsychopharmacology (ECNP) Medicines Chest Initiative: Rationale and promise | Repurposing, Shelved, Discontinued | Efficacy, Strategic reasons |  | Collaborative initiatives, Compound libraries and/or drug-disease databases | CNS | Article | 5 |
| 2008 | Heltzer | Government Cancels Testing of Experimental AIDS Vaccine; Large-Scale Study to Be Scaled Back After Similar Drug Failed; Roche to suspend HIV research, seeing no advances | Suspended, Cancelled | Efficacy |  |  | HIV/AIDS | Article | 5 |
| 2012 | Herschel | Portfolio decisions in early development (Don't throw ouw the baby with the bathwater) |  | Safety issues, Efficacy, Strategic reasons | Access to data |  |  | Expert Opinion/ Editorial | 5 |
| 2016 | Hwang | Failure of investigational drugs in late-stage clinical development and publication of trial results |  | Safety issues, Efficacy, Strategic reasons |  |  |  | Article | - |
| 2018 | Ismail | The potential and benefits of repurposing existing drugs to treat rare muscular dystrophies | Repositioning, Repurposing, Reprofiling, Therapeutic switching, Abandoned |  | Out of scope/ lack of expertise, Patent and IP issues, Access to data, Financing | Collaborative initiatives, Compound libraries and/or drug-disease databases | Duchenne muscular dystrophy | Expert Opinion/ Editorial | 5 |
| 2014 | Jin | Toward better drug repositioning: Prioritizing and integrating existing methods into efficient pipelines | Repositioning |  |  | Use of computational methods/systematic screening methods |  | Review | - |
| 2000 | Johannes | Medicines Co. Gets Federal Approval of Blood-Clot Drug | Abandoned | Efficacy, Strategic reasons |  | Funding structures |  | Press release/ New Article | 5 |
| 2014 | Kadioglu | Contributions from emerging transcriptomics technologies and computational strategies for drug discovery | Repositioning, Reprofiling, Failed |  |  | Compound libraries and/or drug-disease databases, Use of computational methods/systematic screening methods |  | Review | - |
| 2011 | Kaiser | NIH's Secondhand Shop for Tried-and-Tested Drugs | Repurposing, Rescue, Shelved, Abandoned, Failed | Efficacy | Patent and IP issues, Access to data, Testing | Collaborative initiatives |  | Press release/ New Article | 5 |
| 2016 | Katare | Repositioning of Drugs in Cardiometabolic Disorders: Importance and Current Scenario | Repositioning, Shelved | Safety issues, Efficacy | Patent and IP issues, Low ROI potential | Regulatory modifications | Cardiometabolic disorder | Review | - |
| 2018 | Kerrigan | Repurposing old ways of treating sepsis | Repurposing, Failed | Efficacy |  |  | Sepsis | Press release/ New Article | 5 |
| 2017 | Khaladkar | Uncovering novel repositioning opportunities using the Open Targets platform | Repositioning, Repurposing, Failed |  |  | Compound libraries and/or drug-disease databases, Use of computational methods/systematic screening methods | Rare disease | Review | - |
| 2018 | Khanna | Entangled decisions: Knowledge interdependencies and terminations of patented inventions in the pharmaceutical industry |  | Managerial decisions |  |  |  | Article | 5 |
| 2015 | Kim | Drug repositioning approaches for the discovery of new therapeutics for Alzheimer’s disease | Repositioning, Repurposing | Safety issues, Efficacy |  | Collaborative initiatives, Compound libraries and/or drug-disease databases, Use of computational methods/systematic screening methods | Alzheimer's disease | Review | - |
| 2017 | Kresge | Why Big Pharma is Willing to lose fortune seeking Alzheimer’s cure | Abandoned, Failed | Efficacy |  |  | Alzheimer's disease | Article | 5 |
| 2019 | Kuchler | Biogen shares soar on plan to seek Alzheimer's drug approval |  | Efficacy |  |  |  | Press release/ New Article | 5 |
| 2019 | Langreth | Confusing Alzheimer's data leaves big decision in FDA hands |  | Efficacy |  |  |  | Press release/ New Article | 5 |
| 2011 | Loging | Cheminformatic/bioinformatic analysis of large corporate databases: Application to drug repurposing | Repositioning, Repurposing |  | Access to data | Compound libraries and/or drug-disease databases, Use of computational methods/systematic screening methods |  | Article | 5 |
| 2014 | Lotharius | Repositioning: The fast track to new anti-malarial medicines? | Repositioning, Repurposing, Discontinued |  | Access to data, Resources | Compound libraries and/or drug-disease databases | Malaria | Article | 4 |
| 2015 | Mandrioli | Discontinued anxiolytic drugs (2009-2014) | Repositioning, Abandoned,  Discontinued | Safety issues, Efficacy, Strategic reasons | Bias, Costs |  | Anxiety | Article | 5 |
| 2020 | Marrs | Phosphonopeptides Revisited, in an Era of Increasing Antimicrobial Resistance | Revisit, Re-evaluation, Abandoned | Strategic reasons |  |  | Antimicrobials | Article | 4 |
| 2011 | Marusina | The CTSA Pharmaceutical Assets Portal a public private partnership model for drug repositioning | Repositioning, Repurposing | Safety issues, Efficacy, Strategic reasons | Bias, Out of scope/ lack of expertise, Patent and IP issues, Low ROI potential, Liability, Access to data, Costs | Collaborative initiatives, Compound libraries and/or drug-disease databases | Rare disease, Neglected diseases | Article | 4 |
| 2013 | Maximov | The Discovery and Development of Selective Estrogen Modulators (SERMs) for Clinical Practice | Abandoned |  |  |  |  | Review | - |
| 2006 | Mayor | Cancer Charity is to "borrow" candidate drugs shelved by companies |  | Strategic reasons | Low ROI potential, Liability | Collaborative initiatives | Cancer | Press release/ New Article | 5 |
| 2016 | Mehndiratta | Drug repositioning | Repositioning, Repurposing, Reprofiling, Rescue, Retasking, Redirecting | Efficacy | Access to data | Use of computational methods/systematic screening methods | Epilepsy | Review | - |
| 2017 | Mehta | Why do trials for Alzheimer's disease drugs keep failing? A discontinued drug perspective for 2010-2015 |  | Safety issues, Efficacy, Methods Flaws |  |  |  | Expert Opinion | 5 |
| 2015 | Millan | Learning from the past and looking to the future: Emerging perspectives for improving the treatment of psychiatric disorders | Repurposing, Rescue, Discontinued, Unmarketed |  |  | Use of computational methods/systematic screening methods, Other |  | Review | - |
| 2012 | Mullard | Drug repurposing programmes get lift off | Repositioning, Repurposing, Abandoned, Shelved, Deprioritized, Failed | Efficacy, Strategic reasons | Bias, Patent and IP issues, Risk | Collaborative initiatives |  | Article | 5 |
| 2014 | Mullard | Bank tests drug development waters | Deprioritized |  |  | Collaborative initiatives |  | Press release/ New Article | 5 |
| 2013 | Novac | Challenges and opportunities of drug repositioning | Repositioning | Safety issues, Efficacy, Strategic reasons |  | Collaborative initiatives, Extended profiling, Biotech repositioning companies |  | Expert Opinion/ Editorial | 5 |
| 2014 | Naylor | Therapeutic drug repurposing, repositioning and rescue Part 1: Overview | Repositioning, Repurposing, Rescue | Efficacy | Bias, Out of scope/ lack of expertise, Patent and IP issues, Safety | Collaborative initiatives, Compound libraries and/or drug-disease databases, Open-source initiatives | Rare diseases | Review | - |
| 2019 | Neuberger | Renovation as innovation: Is repurposing the future of drug discovery research? | Repositioning, Repurposing |  |  | Collaborative initiatives, Compound libraries and/or drug-disease databases, Grant funding |  | Expert Opinion/ Editorial | 5 |
| 2016 | Nosengo | New tricks for old drugs | Repositioning, Failed |  | Bias, Out of scope/ lack of expertise, Patent and IP issues, Low ROI potential, Access to data, Risk, Lack of transparency | Collaborative initiatives, Compound libraries and/or drug-disease databases |  | Article | 5 |
| 2005 | Offit | Why are pharmaceutical companies gradually abandoning vaccines? |  | Safety issues, Strategic reasons, Effect of Mergers, Product liabilities |  |  |  | Article | 5 |
| 2008 | Opar | Mixed results for disease-modification strategies for Alzheimer's disease | Failed | Efficacy |  |  |  | Review | - |
| 2018 | Ortuso | The Mu.Ta. Lig Chemotheca: A community-populated molecular database for multi-target ligands identification and compound-repurposing | Repurposing, Existing | Pharma-cological profile |  | Compound libraries and/or drug-disease databases |  | Article | 4 |
| 2018 | Parasrampuria | Why Drugs Fail in Late Stages of Development: Case Study Analyses from the Last Decade and Recommendations | Failed | Safety issues, Efficacy, Trial design |  |  |  | Commentary | 5 |
| 2016 | Pisanu | Lithium pharmacogenetics: Where do we stand? | Repurposing |  |  |  |  | Review | - |
| 2019 | Pizzorno | Fighting the Damocles Sword of Infectious Diseases through Drug Repurposing | Repositioning, Repurposing, Shelved | Efficacy | Bias | Use of computational methods/systematic screening methods | Infectious Disease | Expert Opinion/  editorial | 5 |
| 2014 | Plumridge | Drug Makers Tiptoe Back into antibiotic R&D | Abandoned | Strategic reasons |  |  | Bacterial infection | Press release/ New Article | 5 |
| 2019 | Polamreddy | The drug repurposing landscape from 2012 to 2017: Evolution, challenges, and possible solutions | Repositioning, Repurposing, Reprofiling |  | Patent and IP issues, Costs | Collaborative initiatives, Engagement of stakeholders | Rare diseases | Review | - |
| 2012 | Powell | Silence is not the best medicine: Requiring disclosure of clinical trial data for abandoned drugs |  | Safety issues, Strategic reasons | Bias, Patent and IP issues, Access to data | Disclosure of clinical trial information |  | Commentary | 5 |
| 2018 | Pulley | Advocating for mutually beneficial access to shelved compounds | Repositioning, Repurposing, Resurrect | Efficacy | Lack of incentives, Low reporting failures | Collaborative initiatives, Regulatory modifications, Tax incentives, Company readiness and willingness to share |  | Expert Opinion/ Editorial | 5 |
| 2017 | Pulley | Accelerating precision drug development and drug repurposing by leveraging human genetics | Repositioning, Repurposing, Rescue | Efficacy | Out of scope/ lack of expertise, Institutional champion, Cost | Use of computational methods/systematic screening methods |  | Technical report | 5 |
| 2020 | Pulley | Using what we already have: Uncovering new drug repurposing strategies in existing omics data | Repurposing |  |  | Compound libraries and/or drug-disease databases, Use of computational methods/systematic screening methods |  | Review | - |
| 2019 | Pushpakom | Drug repurposing: Progress, challenges, and recommendations | Repositioning, Repurposing, Reprofiling, Re-tasking, Deprioritized, Discontinued | Efficacy | Patent and IP issues, Access to data, Cost, Organizational hurdles, Regulatory considerations | Collaborative initiatives |  | Review | - |
| 2011 | Rogawski | Disclosure of clinical trial results when product development is abandoned |  | Safety issues,  Efficacy,  Strategic reasons | Patent and IP issues, Access to data |  |  | Expert Opinion/ Editorial | 5 |
| 2017 | Roland | AstraZeneca Gives Rejected Drugs a Second Life; Company has outside researchers look for untapped potential in drugs it had shelved, in an attempt to improve R&D output |  | Efficacy,  Strategic reasons | Low ROI potential | Collaborative initiatives | Bacterial infections | Press release/ New Article | 5 |
| 2004 | Rowland | Firms abandoning antibiotics research | Abandoned | Strategic reasons |  |  |  | Press release/ New Article | 5 |
| 2016 | Saito | Drug Development Abandonment Stage for Japanese Pharmaceutical companies |  | Strategic reasons |  |  |  | Article |  |
| 2012 | Sanseau | Use of genome-wide association studies for drug repositioning | Repositioning |  |  | Use of genome-wide association studies |  | Correspondence | 5 |
| 1995 | Schwartz | AIDS Research: Who Drives the train? | Failed |  |  |  | AIDS | Press release/ New Article | 5 |
| 2013 | Sekhon | Repositioning drugs and biologics: Retargeting old/existing drugs for potential new therapeutic applications | Repositioning, Repurposing, Reprofiling, Rescue, Retasking, Therapeutic switching, Failed, Stalled | Safety issues,  Efficacy,  Strategic reasons | Out of scope/ lack of expertise, Patent and IP issues, Access to data, Cost | Collaborative initiatives, Compound libraries and/or drug-disease databases, Use of computational methods/systematic screening methods | Rare and neglected diseases | Article | 5 |
| 2018 | Shahreza | A review of network-based approaches to drug repositioning | Repositioning, Repurposing, Reprofiling, Retasking, Redirecting, Therapeutic switching, Failed |  |  | Use of computational methods/systematic screening methods |  | Review | - |
| 2002 | Silber | Emeryville, Calif. - Based Biotechnology Firm Dampens Forecast for Profit |  | Strategic reasons, Method flaws |  |  |  | Press release/ New Article | 5 |
| 2006 | Simons | Big Pharma's New R&D CENTER: THE TRASH BIN | Repositioning, Failed, Discarded |  |  | Deal/payment structures |  | Press release/ New Article | 5 |
| 2011 | Smith | Repositioned drugs: Integrating intellectual property and regulatory strategies | Repositioning, Repurposing, Abandoned, Shelved |  |  | Patent |  | Viewpoint | 5 |
| 2019 | Southall | Freedom of information act access to an investigational new drug application | Repurposing | Strategic reasons | Access to data |  |  | Viewpoint | 5 |
| 2011 | Swamidass | Mining small-molecule screens to repurpose drugs | Repositioning, Failed, Stalled |  | Access to data | Use of computational methods/systematic screening methods |  | Viewpoint | 5 |
| 2006 | Swiatek | Eli Lilly to revisit its drug scrap bin | Repositioning, Rescrutinized, Deprioritized, Stalled, Unsuccessful |  |  | Collaborative initiatives, Regulatory modifications, Licensing structures |  | Press release/ New Article | 5 |
| 2020 | Talevi | Challenges and opportunities with drug repurposing: Finding strategies to find alternative uses of therapeutics | Repositioning, Repurposing, Reprofiling, Indication expansion, Indication shift, Shelved, Discontinued |  | Out of scope/ lack of expertise, Patent and IP issues, Access to data, Internal procedures | Collaborative initiatives | Rare and neglected conditions | Expert Opinion/ Editorial | 5 |
| 2006 | Tartaglia | Complementary new approaches enable repositioning of failed drug candidates | Repositioning | Safety issues,  Efficacy,  Strategic reasons | Out of scope/ lack of expertise | Use of computational methods/systematic screening methods, Method-of-use patent |  | Expert Opinion/ Editorial | 5 |
| 2016 | Townsend | Reducing the risk of failure: Biomarker-guided trial design |  | Methods flaws |  |  |  | Comment | 5 |
| 2018 | van Wijk | Positive and Negative AIT trials: what makes the difference? | Failed | Efficacy,  Methods flaws |  |  | Allergic rhinitis and asthma | Review | - |
| 2014 | Verhaar | Repurposing Miltefosine for the Treatment of Immune-Mediated Disease | Repurposing, Failed | Efficacy |  |  | Immune-mediated disease, Parasitic disease | Review | - |
| 2012 | Wadman | New Cures sought from old drugs | Repurposing, Resurrecting, Redeploying, Re-examining | Efficacy,  Strategic reasons | Bias, Patent and IP issues, Access to data | Collaborative initiatives | Progeria | Press release/ New Article | 5 |
| 2013 | Wang | Predicting drug-target interactions using restricted Boltzmann Machines | Repositioning, Repurposing, Abandoned |  |  |  |  | Article | 3 |
| 2011 | Wechsler | Who will fund pharmaceutical R&D | Repurposing, Rescue, Abandoned | Safety issues, Strategic reasons |  | Collaborative initiatives |  | Expert Opinion/ Editorial | 5 |
| 2013 | Williams | Discontinued drugs in 2012: Oncology drugs | Failed | Safety issues,  Efficacy |  | Collaborative initiatives |  | Expert Opinion/ Editorial | 5 |
| 2016 | Wurth | Drug-repositioning opportunities for cancer therapy: Novel molecular targets for known compounds | Repositioning, Repurposing, Reinvestigating, Shelved |  | Risk | Use of computational methods/systematic screening methods, Economic support | Cancer | Review | - |
| 2014 |  | World's largest collection of deprioritized pharma compounds opens to researchers |  | Efficacy | Access to data | Collaborative initiatives, Compound libraries and/or drug-disease databases | Neurological disease | Press release/ New Article |  |
| 1997 |  | Business Brief -- AGOURON PHARMACEUTICALS INC.: Pharmaceutical Firm Drops Experimental Cancer Drug |  | Efficacy,  Strategic reasons |  |  |  | Press release/ New Article |  |
| 2012 |  | NIH Initiative to encourage researchers to find new uses for experimental drugs | Repurposing, Failed, Old, Experimental, Existing |  |  | Collaborative initiatives |  | Press release/ New Article |  |
| 2017 |  | Scientists aim to repurpose former experimental cancer therapy to treat muscular dystrophy | Repurposing, Unsuccessful |  |  | Collaborative initiatives, Compound libraries and/or drug-disease databases | Muscular Dystrophy | Press release/ New Article |  |
| 2012 |  | Roche scraps Cholesterol Drug for Lack of benefit | Abandoned | Safety issues,  Efficacy |  |  | Heart disease | Article |  |
| 2012 |  | GSK will reveal more drug information from clinical trials | Abandoned |  |  | Release of clinical trial information |  | Press release/ New Article | 5 |
| 2012 |  | AstraZeneca | Abandoned, Failed | Efficacy |  |  |  | Press release/ New Article | 5 |
| 2007 |  | New Dangers seen in Pfizer's abandoned experimental heart drug | Abandoned | Safety issues,  Efficacy |  |  |  | Press release/ New Article | 5 |
| 2019 |  | Biogen Stock plunges after company halts Alzheimer's Trials | Failed | Efficacy,  Strategic reasons |  |  | Alzheimer's disease | Press release/ New Article | 5 |
